# Gasdermin-D activation by SARS-CoV-2 trigger NET and mediate COVID-19 immunopathology

**DOI:** 10.1101/2022.01.24.22269768

**Authors:** Camila Meirelles Silva, Carlos Wagner S Wanderley, Flavio Protasio Veras, Augusto Veloso Gonçalves, Mikhael Haruo Fernandes Lima, Juliana E. Toller Kawahisa, Giovanni Freitas Gomes, Daniele Carvalho Nascimento, Valter V. Silva Monteiro, Isadora Marques Paiva, Cícero José Luíz Ramos Almeida, Diego Brito Caetité, Juliana da Costa Silva, Maria Isabel Fernandes Lopes, Letícia Pastorelli Bonjorno, Marcela Cavichioli Giannini, Natalia Brasil Amaral, Maíra Nilson Benatti, Luis Eduardo Alves Damasceno, Bruna Manuella Souza Silva, Ayda Henriques Schneider, Icaro Maia Santos Castro, Juan Carlo Santos Silva, Amanda Pereira Vasconcelos, Tiago Tomazini Gonçalves, Sabrina Setembre Batah, Tamara Silva Rodrigues, Victor Ferreira Costa, Marjorie Cornejo Pontelli, Ronaldo B Martins, Timna Varela Martins, Danillo Lucas Alves Espósito, Guilherme Cesar Martelossi Cebinelli, Benedito Antônio Lopes da Fonseca, Luiz Osório Silveira Leiria, Larissa Dias Cunha, Eurico Arruda, Helder I Nakaia, Alexandre Todorovic Fabro, Renê D Oliveira, Dario S Zamboni, Paulo Louzada Junior, Thiago Mattar Cunha, José Carlos Farias Alves Filho, Fernando de Queiroz Cunha

## Abstract

The release of neutrophil extracellular traps (NETs) is associated with inflammation, coagulopathy, and organ damage found in severe cases of COVID-19. However, the molecular mechanisms underlying the release of NETs in COVID-19 remain unclear. Using a single-cell transcriptome analysis we observed that the expression of GSDMD and inflammasome-related genes were increased in neutrophils from COVID-19 patients. Furthermore, high expression of GSDMD was found associated with NETs structures in the lung tissue of COVID-19 patients. The activation of GSDMD in neutrophils requires live SARS-CoV-2 and occurs after neutrophil infection via ACE2 receptors and serine protease TMPRSS2. In a mouse model of SARS-CoV-2 infection, the treatment with GSDMD inhibitor (disulfiram) reduced NETs release and organ damage. These results demonstrated that GSDMD-dependent NETosis plays a critical role in COVID-19 immunopathology, and suggests that GSDMD inhibitors, can be useful to COVID-19 treatment.

**In Brief:** Here, we showed that the activation of the Gasdermin-D (GSDMD) pathway in neutrophils controls NET release during COVID-19. The inhibition of GSDMD with disulfiram, abrogated NET formation reducing lung inflammation and tissue damage. These findings suggest GSDMD as a target for improving the COVID-19 therapy.

## Introduction

NETs are networks composed of extracellular DNA containing histones and cytotoxic enzymes, including myeloperoxidase and elastase, which are released by activated neutrophils as part of their microbicidal arsenal (Brinkmann et al., 2004). However, NETs can also induce disseminated vascular coagulation, when released into the circulation, as well as tissue damage when released in the extravascular space (Knight et al., 2014; Carmona-Rivera et al., 2020; Czaikoski et al., 2016; Papayannopoulos 2018). In this context, NETs have been described as a key mediator of the pathogenesis of various inflammatory conditions, including COVID-19 (Papayannopoulos, 2018; Leppkes et al., 2020; Veras et al., 2020; Ackermann et al., 2021).

We and others demonstrated that patients with severe COVID-19 have an increased number of circulating- and lung-infiltrated neutrophils, which release a large amount of NETs (Veras et al., 2020; Ackermann et al., 2021; Radermecker et al., 2020), mediating lung epithelial damage and disseminated intravascular coagulation. However, the molecular mechanisms involved in NET release during SARS-CoV-2-induced response were not addressed.

Recent works showed that Gasdermin-D (GSDMD) is critical to the release of NETs (Sollberger et al., 2018; Chen et al., 2018). GSDMD cleavage by inflammatory caspases generates two fragments N and C-terminal. The N-terminal (GSDMD-NT) oligomerizes with plasma and nuclear membranes, forming membranes pores that mediate cell death by NETosis (Sollberger et al., 2018; Chen et al., 2018; Kambara et al., 2018; Broz et al., 2020). During sepsis, the genetic deletion or pharmacological inhibition of GSDMD with disulfiram prevented the formation of NETs, protecting mice from organ damage development and increasing survival (Silva et al., 2021). Disulfiram is a drug that inhibits aldehyde dehydrogenase (ALDH) and is used to treat alcoholism (Koppaka et al., 2012). It was demonstrated that disulfiram can act as a potent inhibitor of GSDMD (Hu et al., 2020), and an observational study based on clinical records from the national US Veterans Affairs healthcare system revealed a reduced risk of SARS-CoV-2 infection and deaths in individuals treated with disulfiram (Filmore et al., 2021). Although NETosis is critical for the outcome of COVID-19 (Leppkes et al., 2020; Veras et al., 2020; Ackermann et al., 2021), the mechanisms by which SARS-CoV-2 induced NETs remain unclear.

In the present study, we identified a key role of the GSDMD pathway on NET release during COVID-19. Notably, the inhibition of GSDMD with disulfiram, abrogated NET formation, reducing lung inflammation and tissue damage in a mouse model of COVID-19. These findings indicate GSDMD as a novel potential target for improving the COVID-19 therapeutic strategy.

## Results

### Expression of activated GSDMD is associated with NETosis during COVID-19

To investigate the molecular pathway involved in NETs production in COVID-19, we first performed a single-cell RNA sequencing (scRNA-seq) analysis in public data of bronchoalveolar lavage fluid (BALF) from COVID-19 patients and healthy controls (**Liao et al**., **2020**). Confirming previous findings clustering analysis showed that BALFs of patients with severe COVID-19 contained higher proportions of neutrophils than healthy controls or patients with moderate COVID-19 **(Fig. 1 A)**. The analysis of gene expression in these neutrophils revealed that GSDMD is expressed and most part of these cells also expressed other inflammasome-related genes **(Fig. 1 B)**. We identified neutrophils with three molecular profiles according to the expression of inflammasome genes (*Pycard, Casp4*, and *Casp1*) as indicated by linked points (**Fig. 1 C**). Then, we enrolled 63 hospitalized patients with moderate [n=18, age 63 (±16.58) years, and 55.5% women] and severe [n=45, age 58 (±16.85) years, and 48.8% women] COVID-19. Assisted mechanical ventilation was implemented in 100% of patients with severe COVID-19. The serum levels of CRP, LDH, and fibrin degradation products D-dimers, were found above the normal range, indicating ongoing inflammation, tissue lesions, thrombosis, and subsequent fibrinolysis. The identified causes of death were pneumonia, ARDS, or multi-organ failure. Existing comorbidities are also reported in **Table 1**. Confirming previous findings (Veras et al., 2020), COVID-19 patients showed higher levels of NETs in the airway fluid (**Fig. S1 A**). Notably, the cleaved fraction of GSDMD (GSDMD-NT) was found on these NETs (**Fig. S 1B**). Moreover, the image analysis of lung autopsies from patients who died by COVID-19 showed the presence of NET structure associated with activated GSDMD-NT fraction (**Fig. 1, D-F**). As a control, NETs and GSDMD-NT were not found in the tissues obtained in lung autopsies from patients who died of acute myocardial infarction (**Fig. 1 D**). Furthermore, in lung tissue, it was observed a positive correlation between GSDMD-NT:DAPI colocalization with NETs (MPO:DAPI), confirmed by Pearson’s correlation coefficient analysis (**Fig. 1 G**). Then, we analyzed the expression of GSDMD in blood neutrophils from COVID-19 patients and healthy volunteers. Using confocal microscopy, we found elevated expression of GSDMD accumulated on the neutrophil plasmatic membrane and in typical NETs structures containing extracellular DNA (**Fig. 1, H and I**). The western blot analysis confirmed that neutrophils from COVID-19 patients expressed higher levels of GSDMD-NT when compared to controls (**Fig. 1J**). Additionally, we observed that patients with severe COVID-19 showed elevated serum levels of NETs and GSDMD than heath controls and moderate COVID-19 patients (**Fig. 1, K and L**). A positive correlation was found between serum levels of NETs and GSDMD (**Fig. 1 M**). Thus, these results indicate that GSDMD pathway could be involved in the process of COVID-19-induced NETosis.

**Figure 1.**
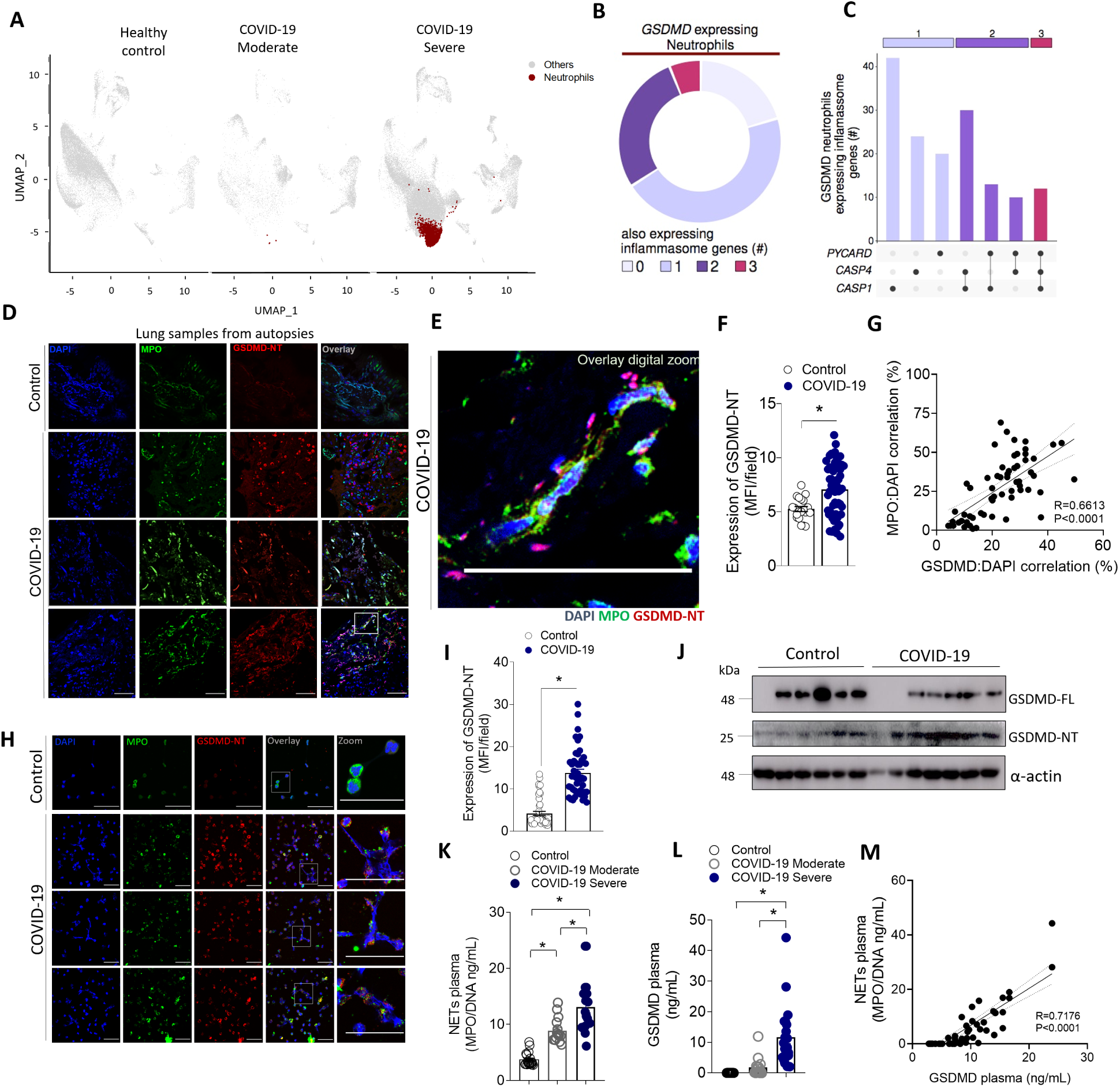
Neutrophils from COVID-19 patients undergoing NETosis express GSDMD. **(A)** Single-cell analysis of BALF from patients with COVID-19 across severity status (Healthy control, Moderate, and Severe). UMAP visualization from gene expression data of 66452 cells, highlighting neutrophil expression cluster in severe COVID-19 patients (red) from bronchoalveolar lavage fluid (BALF) cells. **(B)** Pie chart plot representing the proportion of GSDMD expressing neutrophils. **(C)** UpSet plot showing the intersection of inflammasome genes expressed in neutrophils, including PYCARD, CASP4, and CASP1, derived from COVID-19 severe patients. The point diagram indicates the intersection among the genes and the bar graph shows the number of GSDMD expressing neutrophils in each intersection (y-axis). **(D)** Representative confocal analysis of GSDMD-NT and NETs in the lung tissue sample from autopsies of COVID-19 patients (n=6 or control n=3). Immunostaining for DNA (DAPI, blue), myeloperoxidase (MPO, green), and the GSDMD cleaved fraction (GSDMD-NT, red) are shown. The scale bar indicates 50 μm at 630× magnification. (**E**) Zoomed images of Figure 1D inset white square. **(F)** The expression of GSDMD-NT was quantified by MFI per field. **(G)** Correlation between colocalization GSDMD-NT:DAPI and NETs (MPO:DAPI). **(H)** Representative confocal analysis of GSDMD and NETs in the blood neutrophils isolated from COVID-19 patients (n=5) or controls (n=5). Cells were stained for DNA (DAPI, blue), MPO (green), and GSDMD-NT (red). Scale bar indicates 50 μm, 4× digital zoom was performed in the inset white square. **(I)** Expression of GSDMD-NT was quantified by MFI per field. **(J)** Expression of full-length GSDMD (GSDMD-FL) and active GSDMD (GSDMD-NT) by Western blot. **(K-L)** Circulating amounts of MPO/DNA-NETs and GSDMD from plasma of patients with moderate COVID-19 (n=15) severe COVID-19 (n=21), and healthycontrols (n=20). **(M)** Correlation between plasmatic levels of MPO/DNA-NETs and GSDMD. The data are expressed as mean ± SEM (*p<0.05; t-test in F and I, Pearson’s correlation in G and M, one-way ANOVA followed by Tukey’s in K and L).

**Table 1.** Clinical and laboratory characteristics of COVID-19 patients.

| **Demographics and clinical characteristics of COVID-19 patients** |  |  |  |
| --- | --- | --- | --- |
| **Severity** | Moderate | Severe | P value |
| **Number** | 18 | 45 |  |
| **Age** | 63.29 ± 16.58 | 58.62 ± 16.85 | ns<sup>1</sup> |
| **Female** | 10 (55.55%) | 22 (48.88%) | ns |
| **Respiratory status** |  |  |  |
| **Mechanical ventilation** | 0 | 45 (100%) |  |
| **Nasal-cannula oxygen<sup>Ω</sup>** | 18 (100%) | 0 |  |
| **Room air** | 0 | 0 |  |
| **Outcome** |  |  |  |
| **Secondary infection** | 0 | 12 (26.66%) | 0.013 |
| **Deaths** | 6 (33.33%) | 13 (28.88%) | ns |
| **Clinical - Comorbidities** |  |  |  |
| **Diabetes mellitus** | 6 (33.33%) | 6 (13.33%) | ns |
| **Cardiopathy** | 3 (16.66%) | 5 (11.11%) | ns |
| **Nephropathy** | 1 (5.55%) | 7 (15.55%) | ns |
| **Pneumopathy** | 4 (22.22%) | 6 (13.33%) | ns |
| **Autoimmune diseases** | 1 (5.55%) | 2 (4.44%) | ns |
| **Cancer** | 2 (11.11%) | 2 (4.44%) | ns |
| **Stroke** | 2 (11.11%) | 1 (2.22%) | ns |
| **Obesity** | 5 (27.77%) | 21 (48.88%) | ns |
| **Arterial hypertension** | 9 (50.0%) | 23 (51.11%) | ns |
| **Immunodeficiency** | 2 (11.11%) | 3 (6.66%) | ns |
| **Smoking** | 5 (27.77%) | 9 (20.0%) | ns |
| **Laboratorial Findings** |  |  |  |
| **CRP (mg/dL)\*** | 9.90 ± 6.18 | 12.21 ± 10.13 | ns |
| **D-Dimers (μg/mL)\*\*** | 1.48 ± 1.61 | 3.30 ± 3.55 | 0.007 |
| **LDH (U/L)#** | 428.89 ± 205.23 | 746.60 ± 429.50 | 0.002 |
| **Ferritin (ng/mL)&** | 911.01 ± 712.94 | 1109.47 ± 1547.36 | ns |
| **Neutrophils x10<sup>3</sup>/μL** | 6.0 ± 4.2 | 7.5 ± 3.6 | 0.02 |
| **Lymphocytes x10<sup>3</sup>/μL** | 1.4 ± 0.8 | 1.9 ± 1.5 | ns |
| **Neutrophil/lymphocytes ratio\*** | 4.2 ± 5.25 | 3.9 ± 2.4 | ns |
| **Platelets x10<sup>3</sup>/μL** | 242.7 ± 103.8 | 287.3 ± 121.1 | ns |

<sup>Ω</sup> The patients were classified as moderate at the time of hospital admission, they received nasal oxygen supplementation during hospitalization, but none required mechanical ventilation
<sup>1</sup>ns, not significant
\*CRP, C-reactive protein (normal value < 0.5 mg/dl);
\*\*D-dimers (normal value <0.5 μg/ml);
#LDH, lactic dehydrogenase (normal range: 120–246 U/liter);

### GSDMD is required for NETs release by neutrophils from COVID-19 patients

Recent studies showed that disulfiram potently inhibits GSDMD (Hu et al., 2020). To investigate the potential role of GSDMD on NETs release during COVID-19, we added disulfiram on cell cultures of neutrophils from COVID-19 patients. We found that disulfiram inhibited the release of NETs in a concentration-dependent manner and the expression GSDMD-NT **(Fig. 2, A-C)**. Importantly, the treatment with disulfiram also abrogated the activation of GSDMD and the release of NETs by SARS-CoV-2 infection in neutrophils **(Fig. 2, D-F)**. Of note, disulfiram, at the used concentrations, did not inhibit the viral replication **(Fig S 2)**. These results suggest that GSDMD is involved in the release of NETs triggered by SARS-CoV-2.

**Figure 2.**
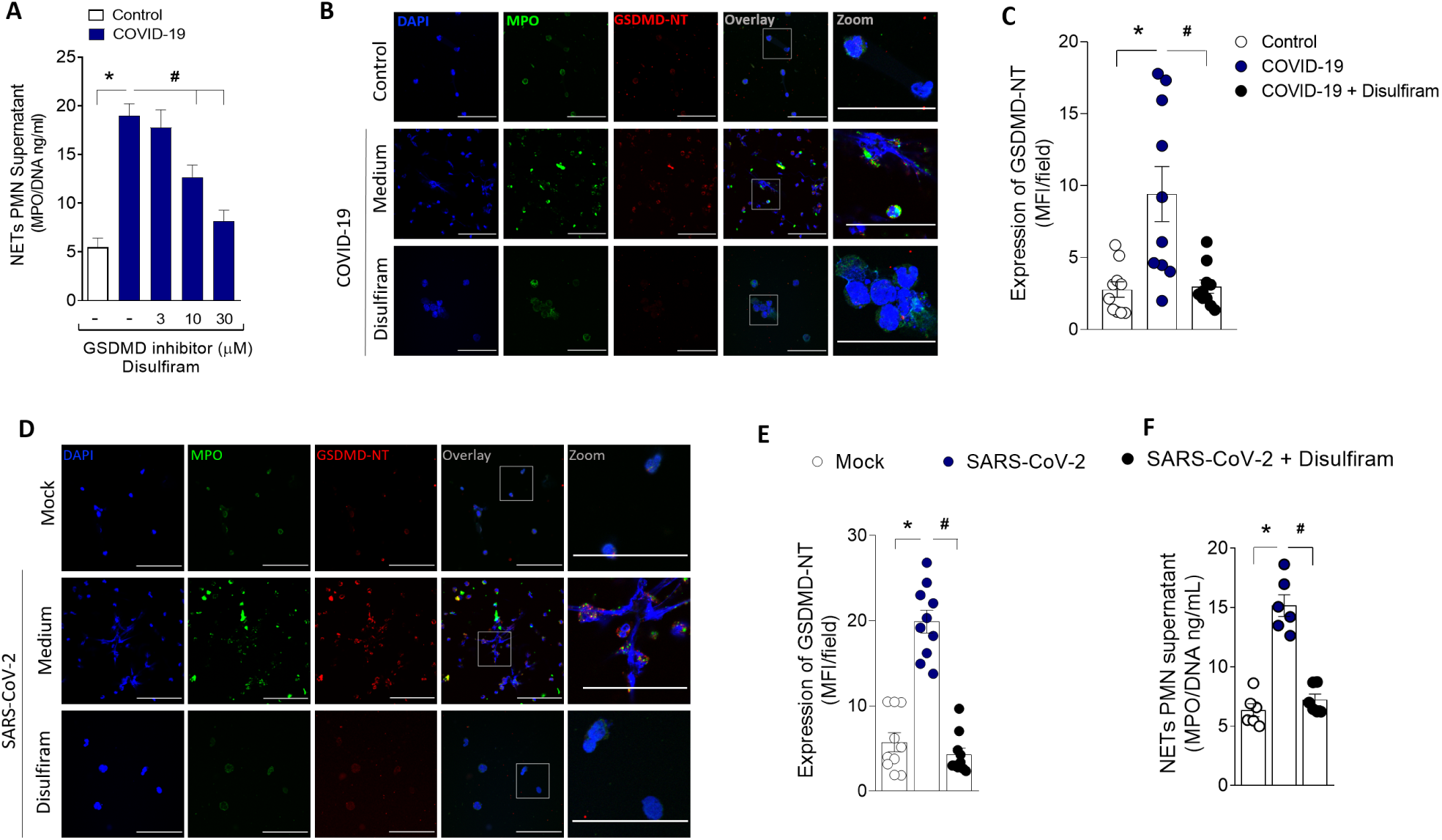
GSDMD activation during COVID-19 mediates NETs formation. Human neutrophils were isolated from healthy control (n=12) and COVID-19 (n=15) patients. Cells were treated with disulfiram (3, 10, and 30 μM) and cultured for 4 h at 37°C. **(A)** The concentrations of MPO/DNA-NETs in the supernatants were determined using the picogreen test. **(B)** Representative immunostaining images for DNA (DAPI, blue), myeloperoxidase (MPO, green), and the GSDMD cleaved fraction (GSDMD-NT, red) are shown. The scale bar indicates 50 μm at 630× magnification. 4× digital zoom was performed in the inset white square. **(C)** GSDMD-NT expression was quantified by MFI per field. (**D**) Human neutrophils were isolated from healthy control (n = 6). Cells were treated with disulfiram (30 uM). After 1 h the cells were incubated with SARS-CoV-2 or Mock (virus control) and cultured for 4 h at 37°C. Representative immunostaining images for DNA (DAPI, blue), myeloperoxidase (MPO, green), and the GSDMD cleaved fraction (GSDMD-NT, red) are shown. The scale bar indicates 50 μm at 630× magnification. 4× digital zoom was performed in the inset white square. **(E)** GSDMD-NT expression was quantified by MFI per field. **(F)** The concentrations of MPO/DNA-NETs in the supernatants were determined using the picogreen test. The data are expressed as mean ± SEM (*or ^#^ p<0.05; one-way ANOVAfollowed by Tukey’s test in A, C, E, and F).

### The cleavage of GSDMD depends on neutrophil infection by SARS-CoV-2

We previously demonstrated that SARS-CoV-2 is able to directly induce NET release by human neutrophils (Veras et al., 2020). Thus, considering that SARS-CoV-2 employs ACE2 and TMPRSS2 for host cell entry (Hoffmann et al., 2020), we investigated whether GSDMD activation in neutrophils is induced by SARS-CoV-2 via ACE2–TMPRSS2 axis during NETosis process. Neutrophils incubation with the inactivated SARS-CoV-2 show neither GSDMD activation nor NETs release. Moreover, treatment of isolated neutrophils with a neutralizing anti-hACE2 antibody (αACE2) or camostat, an inhibitor of TMPRSS2, abrogated SARS-CoV-2-induced GSDMD activation and NETs release. Then, we treated SARS-CoV-2 infected neutrophils with tenofovir disoproxil fumarate (TDF), an antiretroviral that reduces SARS-CoV-2 replication through the inhibition of RNA polymerase (Clososki et al., 2020). Remarkably, TDF also reduced the GSDMD cleavage and release of NETs by neutrophils incubated with SARS-CoV-2 (**Fig. 3, A-C**). These results indicate that SARS-CoV-2 infection and replication activates GSDMD in neutrophils to release NETs. The lack of cleaved GSDMD and NET release in the culture with inactivated SARS-CoV-2 confirms that active viral infection might be necessary to trigger GSDMD-dependent NETosis.

**Figure 3.**
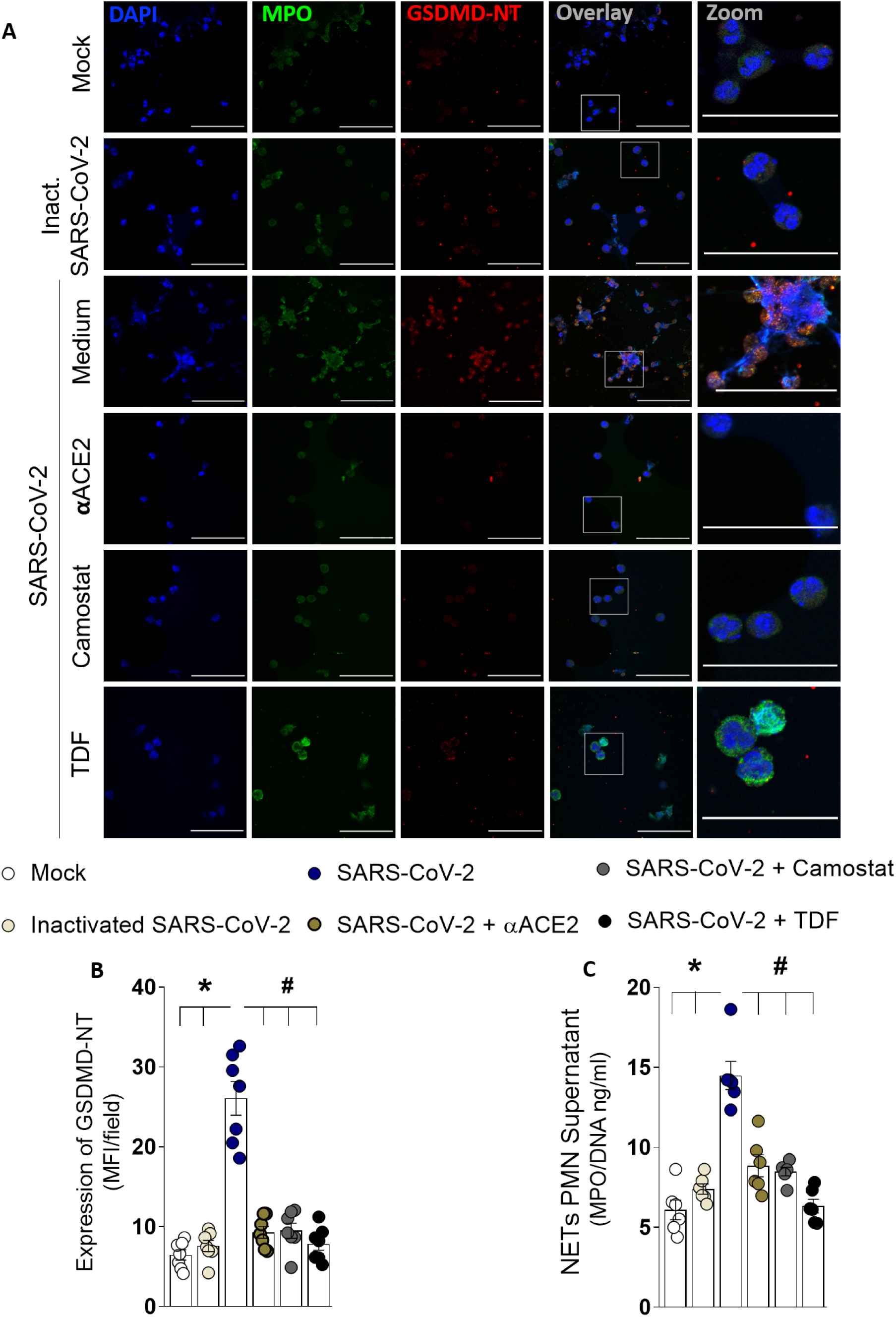
The GSDMD-dependent NETosis is triggered by SARS-CoV-2 directly. Human neutrophils were isolated from healthy control (n=7). Cells were treated with a neutralizing anti-hACE2 antibody (αACE2, 0.5 μg/ml), an inhibitor of the serine protease TMPRSS2 (camostat, 10 μM), or an antiretroviral that reduces SARS-CoV-2 replication through the inhibition of RNA polymerase - tenofovir disoproxil fumarate (TDF; 10 μM). After 1 h the cells were incubated with SARS-CoV-2, or virus control (inactivated SARS-CoV-2 or Mock) and cultured for 4 h at 37°C. **(A)** Representative immunostaining images for DNA (DAPI, blue), myeloperoxidase (MPO, green), and the GSDMD cleaved fraction (GSDMD-NT,red) are shown. The scale bar indicates 50 μm at 630× magnification. 4× digital zoom was performed in the inset white square. **(B)** GSDMD-NT expression was quantified by MFI per field. **(C)** The concentrations of MPO/DNA-NETs in the supernatants were determined using the picogreen test. The data are expressedas mean ± SEM (*or ^#^ p<0.05, one-way ANOVA followed by Tukey’s test in B and C).

### Inflammasome/GSDMD pathway is highly expressed in blood neutrophils from COVID-19 patients

It is well established that inflammasome activation by canonical (caspase-1), or non-canonical pathway (caspase-4) cleaves and activates GSDMD (Chen et al., 2018; Shi et al., 2015).Considering these findings, we investigated whether these pathways are involved in GSDMD activation in COVID-19 patients. Confirming single-cell transcriptome data (**Fig 1 C**), neutrophils from COVID-19 patients also showed increased expression of active caspases-1 and caspase-4 (**Fig. 4 A)**. Also, we observed, that GSDMD activation and NETs production were abrogated in SARS-CoV-2-infected neutrophils from healthy individuals after treatment with inhibitors of caspase-1 (Ac-YVAD-CHO) or pan-caspases (Z-VAD-FMK) **(Fig. 4, B-D)**. Neutrophils from COVID-19 patients also showed increased expression of RIG-I (**Fig. S 3)**, a viral sensor involved in the recognized RNA virus, which is implicated in inflammasome activation (Elion et al., 2018). These results indicate that the inflammasome is involved with the cleavage of GSDMD and NETs release triggered by SARS-CoV-2.

**Figure 4.**
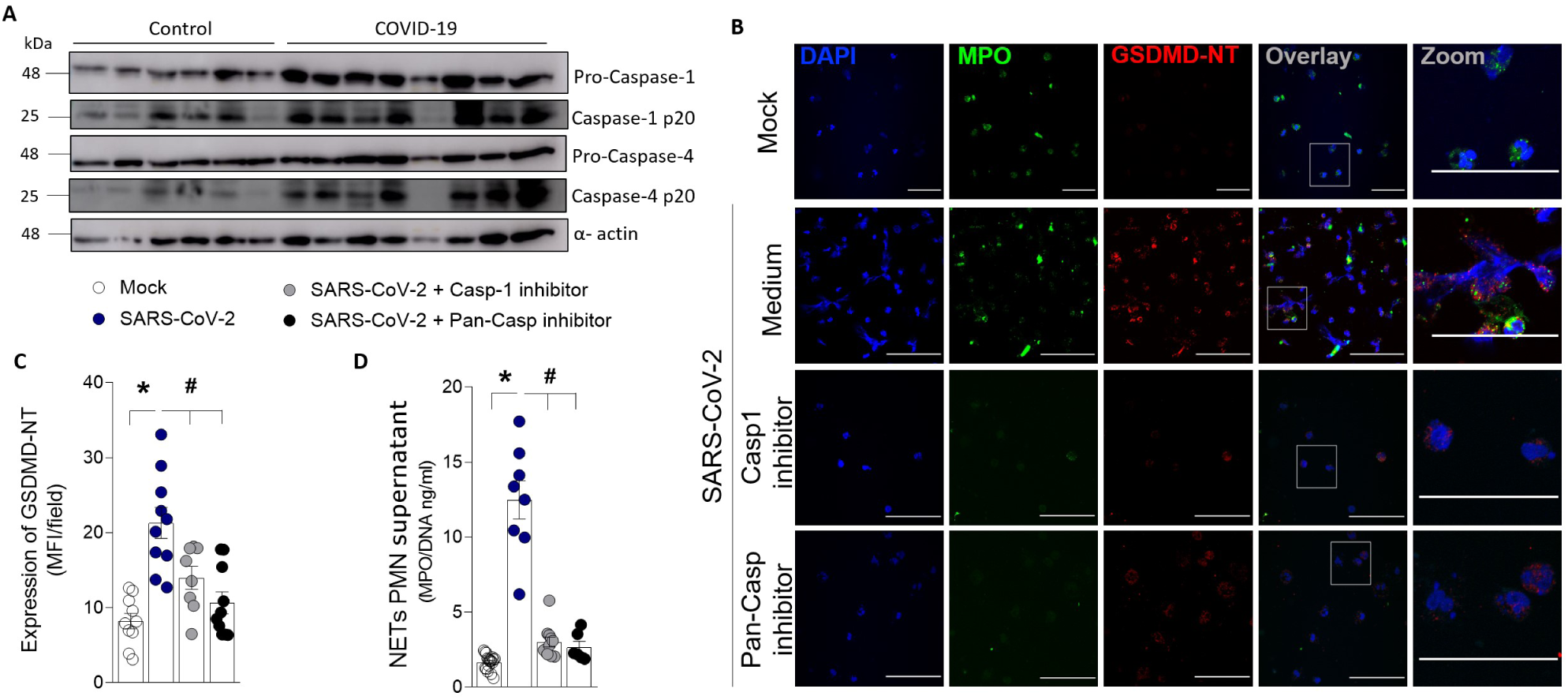
Inflammatory caspases mediate GSDMD cleavage and NETs formation after SARS-Cov-2 neutrophil infection. Neutrophils were isolated from healthy controls (n=6) and COVID-19 patients (n=8). **(A)** The neutrophil lysates were harvested for immunoblot analysis of pro-caspase-1, pro-caspase-4, and their cleaved fraction caspase-1-p20 and caspase-4-p20. The α-actin was used as a loading control. **(B)** Human neutrophils were isolated from healthy controls (n=8). Cells were treated with caspase-1 inhibitor (Ac-YVAD-CHO, 25uM) or pan-caspase inhibitor (Z-VAD-FMK, 50uM). After 1 h the cells were incubated with SARS-CoV-2 or Mock (virus control) and cultured for 4 h at 37°C. Representative immunostaining images for DNA (DAPI, blue), myeloperoxidase (MPO, green), and the GSDMD cleaved fraction (GSDMD-NT, red) are shown. The scale bar indicates 50 μm at 630× magnification. 4× digital zoom was performed in the inset white square. **(C)** GSDMD-NT expression was quantified by MFI per field. **(D)** The concentrations of MPO/DNA-NETs in the supernatants were determined using the picogreen test. The data are expressed as mean ± SEM (*or ^#^ p<0.05, one-way ANOVA followed by Tukey’s test in C and D).

### Epithelial and endothelial cell death elicited by SARS-CoV-2–induced NETs requires GSDMD

In several pathological conditions, the release of NETs is associated with tissue damage (Knight et al., 2014; Carmona-Rivera et al., 2020; Czaikoski et al., 2016; Papayannopoulos 2018; Veras et al., 2020). COVID-19 is characterized by extensive tissue damage, mainly in the lung (Veras et al., 2020; Radermecker et al., 2020, Zeng et al., 2021). Therefore, we investigate whether inhibition of GSDMD prevents NET-induced cell damage. To this end, neutrophils from the blood of healthy controls were treated with disulfiram incubated with SARS-CoV-2 for 1 h, the cells were washed twice, and then co-cultured with a human alveolar basal epithelial cell line (A549) or endothelial cell line (HUVEC) for 24 h. Cell viability was determined (viability dye^+^ cells) by flow cytometry. We found that SARS-CoV-2–activated neutrophils reduced the viability of A549 and HUVEC cells compared with non-activated neutrophils. Importantly, disulfiram treatment reduced the cell death induced by SARS-CoV-2– activated neutrophils (**Fig. 5, A-D and Fig. S 4**). These results indicate that the GSDMD inhibition can prevent tissue damage mediated by SARS-CoV-2–induced NETs.

**Figure 5.**
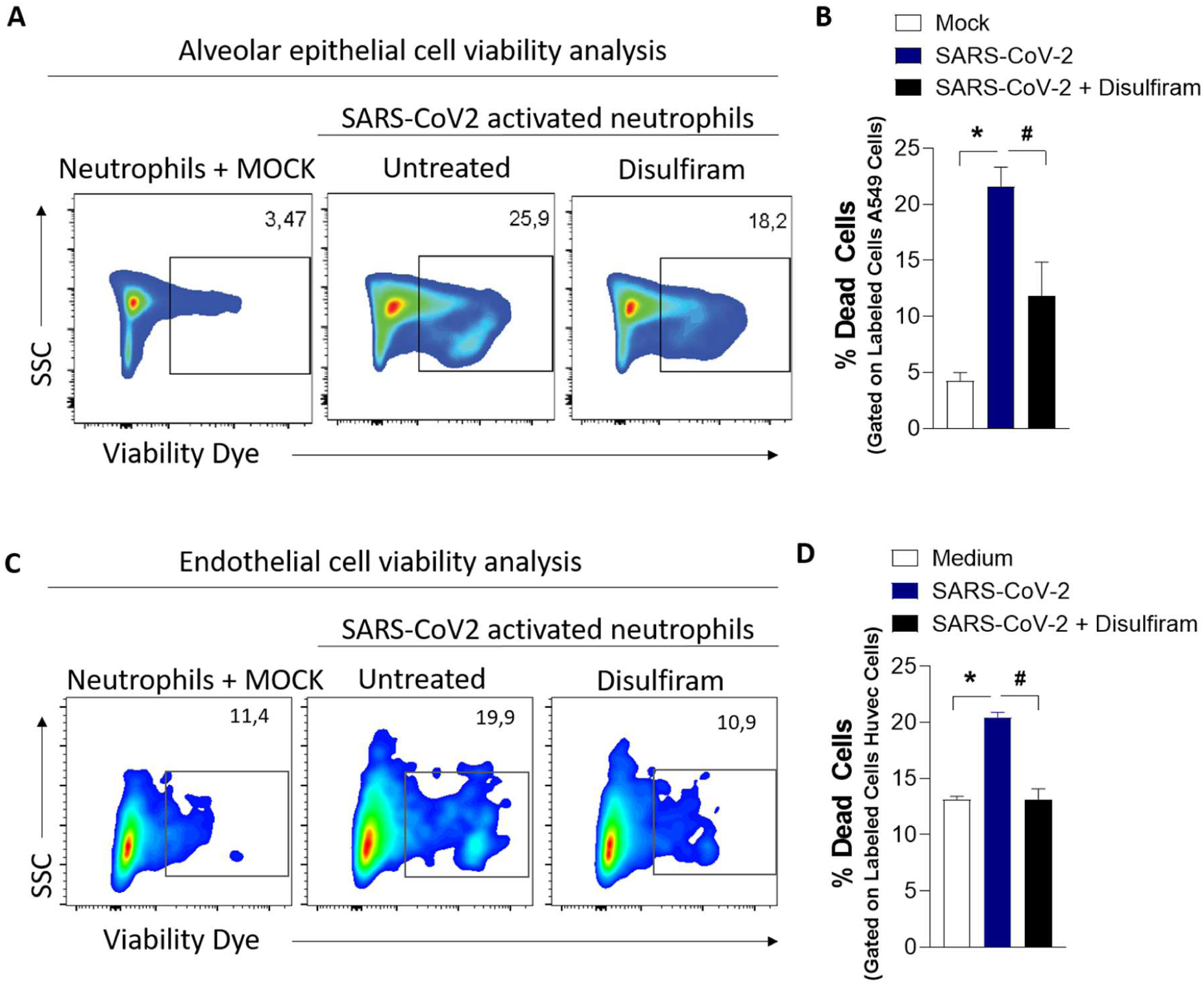
GSDMD inhibition prevents cell damage induced by NETs associated with SARS-CoV-2 infection. Blood isolated neutrophils (1 × 10^6^ cells) from healthy donors, pretreated, or not, with disulfiram (30 μM) were incubated, or not, with SARS-CoV-2 (n=6). After 1 h, these neutrophils were washed twice and co-cultured with lung epithelial cells (A549, 2 × 10^5^ cells) or endothelial cells (HUVEC, 2 × 10^5^ cells) previously stained with viability dye for 24 h at 37°C. **(A)** Representative dot plots of FACS analysis for viability dye+ cells. **(B)** Frequency of viability dye+ A549 cells. **(C)** Representative dot plots of FACS analysis of viability dye+ HUVEC. **(E)** Frequency of viability dye+ HUVEC cells. Data are representative of at least two independent experiments and are shown as mean ± SEM (*or ^#^ p<0.05, one-way ANOVA followed by Tukey’s test in B and D).

### Disulfiram prevents NETs release and organ dysfunction in a COVID-19 experimental model

To investigate the importance of GSDMD-dependent NETosis to COVID-19 immunopathology, we infected K18 hACE2 transgenic mice with SARS-CoV-2 and treated them with disulfiram. According to Oladunni et al. 2020, mice submitted to SARS-CoV infection showed a dramatic reduction in the overall survival curves. In a preliminary experiment, we confirmed these results. Therefore, we perform euthanasia on day 5 after the virus inoculation to obtain tissue and blood samples for analysis. We found that disulfiram treatment reduced the circulating levels of NETs after SARS-CoV-2 infection compared to the group treated with vehicle **(Fig. 6 A)**. Additionally, the levels of inflammatory cytokines IL-6, IL-1β, and CXCL1/KC, but not TNF-α, in lung tissue were also mitigated by treatment with disulfiram **(Fig. 6, B-E)**. Histological analysis of the lung tissue from SARS-CoV-2 infected mice showed a septal thickening by intense neutrophil infiltration with alveolar-capillary barrier damage. At higher magnification, we also observed the parenchymal lung remodeling with architectural distortion and an intense inflammatory cells infiltration with damage of pneumocytes and endothelial cells of the alveolar septa. Importantly, the treatment of infected mice with disulfiram reduced these inflammatory events, avoiding alveolar septal thickening and preserving the tissue histoarchitecture **(Fig. 6 F)**. Furthermore, the confocal microscopy analysis of lung tissues showed that disulfiram treatment markedly reduced the GSDMD-NT expression after SARS-CoV-2 infection **(Fig. 6, G and H)**. Although lung injuries are a hallmark of COVID-19, evidence has shown that other organs are also affected (Zhang et al., 2020; Shi et al., 2020). Thus, we analyze the protective effect of disulfiram in other tissues. We observed that the heart of animals infected with SARS-CoV-2 showed diffuse and sparse cardiac interstitial inflammatory infiltrate, with perivascular accentuation **(Fig. S 5A)**. In the kidney tissue, the SARS-CoV-2 infection-induced ischemic tubulointerstitial nephritis, mimicking acute tubular necrosis, which was associated with cell glomerulitis **(Fig. S 5B)**. In the liver of infected mice, central-portal necroinflammatory hepatitis and spillover with piecemeal necrosis were found **(Fig. S 5C)**. The treatment with disulfiram promoted the preservation of tissue architecture, reduced the inflammatory infiltrate, and attenuated tissue damage. Collectively, these findings demonstrate that pharmacological inhibition of GSDMD with disulfiram prevents NETs release and organ dysfunction and can be used to improve the COVID-19 treatment.

**Figure 6.**
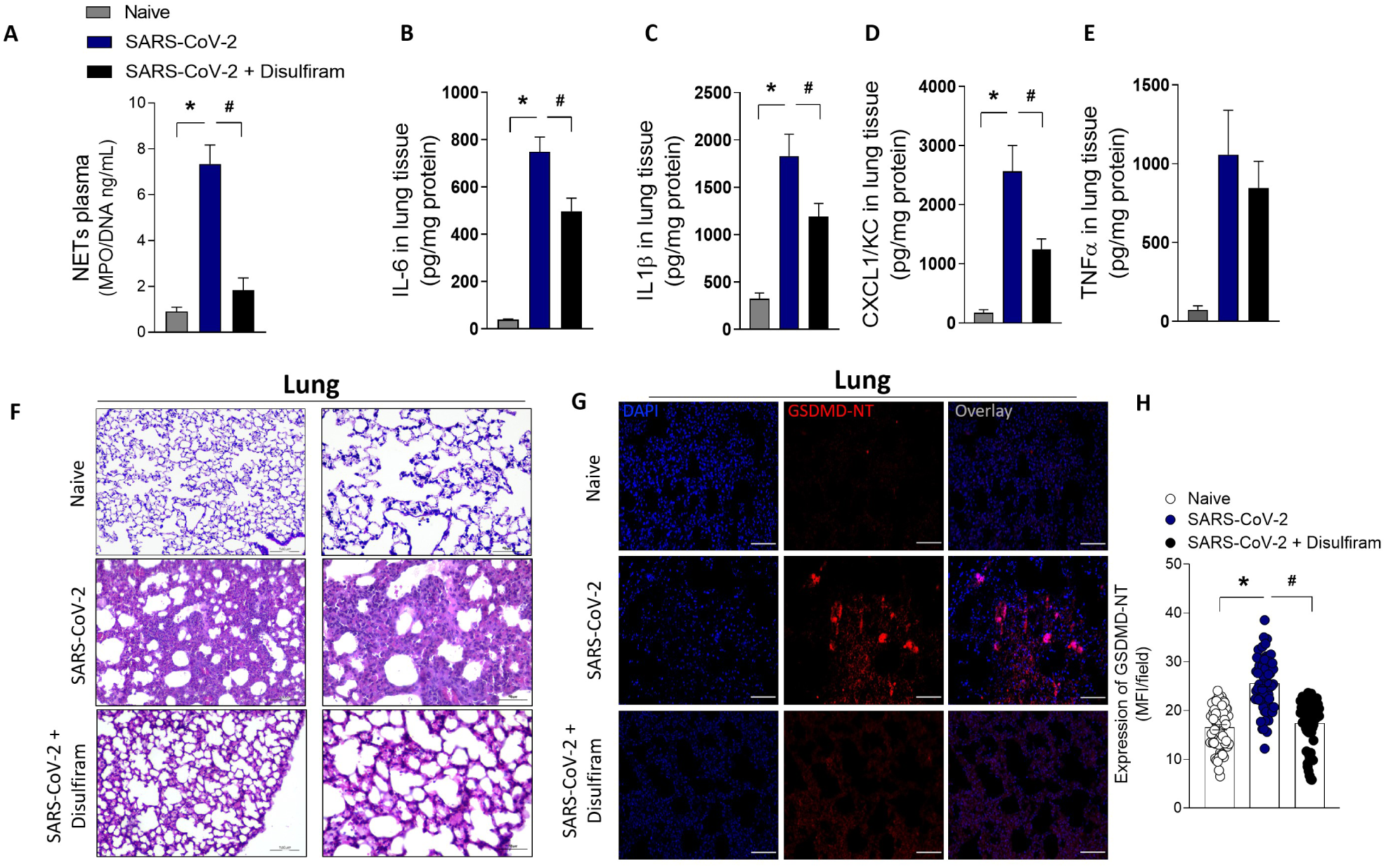
Pharmacological inhibition of GSDMD prevents NET release, lung inflammation, and organ damage in a mouse model of COVID-19. ACE-2 humanized mice were infected with SARS-CoV-2 and after 24 h mice were treated with disulfiram (50 mg/kg, i.p. 1x per day, during 5 days) or vehicle. **(A)** The MPO/DNA-NET concentration in the plasma was determined 5 days post-SARS-CoV-2 infection. **(B-E)** The levels of inflammatory cytokines (IL-6, IL-1β, CXCL-1/KC, and TNF-α) in lung tissue were measured by ELISA 5 days post-SARS-CoV-2 infection. **(F)** Representative images of the histological staining of the lung sections performed 5 days post-SARS-CoV-2 infection are shown at 200×magnification and at 400× magnification. **(G)** Representative confocal analysis of GSDMD-NT and NETs in the lung tissue sample. Immunostaining for DNA (DAPI,blue) and the GSDMD cleaved fraction (GSDMD-NT, red) are shown. The scale bar indicates 50 μm at 630× magnification. **(H)** GSDMD-NT expression was quantified by MFI per field. The data are expressed as means ± SEM (*or ^#^ p<0.05; one-way ANOVA followed by Tukey’s test in A-E and H). The data are representative of at least two independent experiments, each including 5-7 animals per group.

## Discussion

We and others have identified NETs as potential drivers of COVID-19 severity (Leppkes et al., 2020; Veras et al., 2020; Ackermann et al., 2021; Radermecker et al., 2020). The massive release of NETs is associated with systemic inflammation, organ damage, and thrombosis that is found in severe cases of COVID-19 (Leppkes et al., 2020; Veras et al., 2020; Ackermann et al., 2021; Radermecker et al., 2020). Remarkably, we reported that SARS-CoV-2 can directly stimulate human neutrophils to release NETs (Veras et al., 2020). However, how SARS-CoV-2 leads to the release of NETs is still unclear. In the present study, we reveal that the virus that causes COVID-19 directly triggers NET release by a GSDMD pathway-dependent manner. We found that GSDMD is expressed in lung tissue of patients with severe COVID-19 in association with NETs structures. Furthermore, the release of NETs by neutrophils infected with SARS-CoV-2 or isolated from patients with severe COVID-19 was inhibited with GSDMD inhibitor, disulfiram. Moreover, in a mouse model of COVID-19, the treatment with disulfiram abrogated NETs release and reduced organ damage.

A wide array of stimuli can trigger NET release. NETs production can be stimulated by toll-like receptors (TLRs) agonists, inflammasome assembling activators, or by immune complexes via Fc-receptors (Papayannopoulos et al., 2018; Ackermann et al., 2021; Chen et al., 2018). Previous findings demonstrated that a range of bacteria and viruses, including immunodeficiency virus-1 (HIV-1), chikungunya, respiratory syncytial virus (RSV) (Saitoh et al., 2012; Funchal et al., 2015; Hiroki et al., 2020), are able to induce NETs through TLRs and ROS pathways. Although we do not exclude the participation of TLRs and ROS production, especially in the case of secondary infection associated with COVID-19, we are showing that SARS-CoV-2 induced NETs release via inflammasome/GSDMD activation. The activation of this pathway depends on the virus entry into the cell via ACE2 and TMPRSS2 and also effective viral replication. Previous findings demonstrated that SARS-CoV-2 activates caspase-1 in monocytes inducing the production of inflammatory cytokines (Rodrigues., 2020). According to these observations, we demonstrated that after entering and replicating into the neutrophils SARS-CoV-2 activates inflammasome/caspases1/4, which in turn cleaves and activates GSDMD to finally mediate NETs release.

A possible link connecting the activation of caspases by SARS-CoV-2 relies on mechanisms triggered by RNAs sensors. Previous reports indicate that the intracellular RNA sensor RIG-I is involved in the recognition of SARS-CoV-2 (Yamada et al., 2021). The RIG-I activation assembles caspase-1-activating inflammasome complexes and could induce GSDMD cleavage (Elion et al., 2018; Rintahaka et al., 2021). In this context, we observed an increase of RIG-1 expression in neutrophils from COVID-19 patients. Then, the possibility of RIG-I being involved in GSDMD activation via caspases/NLRP3 during SARS-CoV-2 infection deserves future investigation. However, it is important to mention that all findings described here pointed out GSDMD as the final signal for the NETosis induced by SARS-CoV-2. Therefore, we propose GSDMD targeting to ameliorate COVID-19 therapy.

Disulfiram is a drug approved for the treatment of alcohol dependence for its inhibitory effect on aldehyde dehydrogenase (ALDH) (Koppaka et al., 2012; Wright and Moore, 1990). However, a study showed that disulfiram at nanomolar concentration covalently binds and modifies human/mouse Cys191/Cys192 in GSDMD inhibiting its pore-forming function (Hu et al., 2020). Furthermore, we recently demonstrated that inhibition of GSDMD by disulfiram prevents neutrophil death by NETosis (Silva et al., 2021). Considering this finding, we tested the effect of disulfiram on the COVID-19 experimental model. We use K18hACE2 mice with humanized ACE2. K18 hACE2 transgenic mice succumbed to SARS-CoV-2 infection by day 6, with virus detected in lung airway epithelium and brain (Oladunni et al., 2020). We reproduced these data in our conditions at the dose of 2×10^4^ PFU and all experiments were performed with this viral load. Considering that we perform daily treatment starting 24h after the virus inoculation, and on day 5 mice were euthanized to collect samples. This window of therapy with disulfiram was sufficient to reduce NETs concentration, cytokine storm, and attenuate important tissues damages in several organs. Similarly, we observed that the inhibition of GSDMD by disulfiram in the sepsis model efficiently abrogates NETosis, systemic inflammation, and vital organs dysfunction, improving mice survival (Silva et al., 2021). Of note, the course of the disease in the mouse model is different when compared to humans, in which the symptoms are observed on 5–6 days after the infection and maintained for around 14 days (Zhou et a., 2020).

Although the effect of GSDMD inhibition by disulfiram may be clearly associated with the reduction of NETs, we do not exclude its effect in other cells, such as macrophages, blocking the release of inflammatory cytokines, as observed in the lung tissue of infected mice treated with disulfiram.

Taken together, our findings demonstrate that GSDMD plays a critical role in the generation of NETs and organ damage induced by SARS-CoV-2 infection.Therefore, the pharmacological inhibition of GSDMD, as with disulfiram, represents a potential strategy to improve the treatment of COVID-19.

## Material and methods

### Analysis of single-cell RNA-seq data

We analyzed single-cell transcriptomic data from bronchoalveolar lavage fluid (BALF) from patients with varying severity of COVID-19 disease and their respective healthy controls from a previously published cohort (Liao et al., 2020). The dataset generated by authors is publicly available at https://covid19-balf.cells.ucsc.edu/. Basically, the dataset was downloaded and the RDS file was imported into R environment version v4.0.5. UMAP plots were generated using Seurat v4 (Hao et al, 2021). The overlap between gene lists was performed using the UpSetR package (Lex et al, 2014).

### Patients

The inclusion criteria were individuals hospitalized with moderate or severe forms of COVID-19 diagnosed by RT-PCR in nasopharyngeal swab specimens and lung computed tomography scan involvement compatible with COVID-19 pneumonia; older than 18 years; body weight >50 kg; normal levels of serum Ca2+ and K+; QT interval normal levels of serum Ca2+ and K+; QT interval < 450 ms at 12 derivations electrocardiogram (according to the Bazett formula) and negative serum or urinary β-HCG if a woman under 50. The exclusion criteria were defined as diarrhea resulting in dehydration; pregnancy or lactation; metastatic cancer or immunosuppressive chemotherapy and inability to understand the consent form (Lopes et al., 2021). The moderate form was defined in patients with fever, dyspnea, imaging findings of pneumonia but with SatO2>94% on the first day of admission. The severe form was defined in those patients with the same findings of moderate form plus respiratory rate ≥ 30 times per minute or SatO2 ≤ 93% and all of them required mechanical ventilation. Both forms were established on the first day of hospitalization (Wu et al., 2020; Jin et al., 2020). Importantly, the samples were collected on the day of admission. Although the patients were classified as moderate at the time of hospital admission, they received nasal oxygen supplementation during hospitalization, but none required mechanical ventilation. Peripheral blood samples and airway fluid were collected from 63 and 11 confirmed individuals hospitalized with COVID-19, respectively. In the present study, the SARS-CoV-2-negative control group (blood n=20, and airway lavage n=8) was designed to include matched subjects of older age and chronic non-communicable diseases (age, 40.57 ± 15.29; 24% female). The clinical features of the subjects are detailed in **Table 1**. The study was approved by the National Ethics Committee, Brazil (CONEP, CAAE: 30248420.9.0000.5440). Written informed consent was obtained from recruited patients. Lung tissue from 10 patients with COVID-19 was autopsied with the minimally invasive ultrasound-guided approach. The Hospital das Clínicas da Faculdade de Medicina da Universidade de São Paulo (HC-FMUSP) autopsy study was approved under protocol number #3951.904 by HC-FMUSP Ethics Committee and performed on the Autopsy Room Imaging Platform (https://pisa.hc.fm.usp.br/). Lung tissue samples were used for immunostaining previously described (Veras et al., 2020; Duarte-neto et al., 2019). We used, as non-COVID-19 control, lung samples from the autopsy of two patients with acute myocardial infarction under familial consent.

### Plasma and neutrophils isolation

Human circulating neutrophils from patients and healthy controls were isolated using Percoll density gradients. Briefly, peripheral blood samples were collected by venipuncture and centrifuged at 450 x g for plasma separation. The blood cells were then resuspended in Hank’s balanced salt solution (Corning; cat. 21-022-CV), and the neutrophil population was isolated by Percoll (GE Healthcare; cat. 17-5445-01) density gradient (72%, 63%, 54%, and 45%). Isolated neutrophils were resuspended in RPMI 1640 (Corning; cat. 15-040-CVR) supplemented with 0.5% BSA. The neutrophil purity was >95% was determined by Rosenfeld-colored Cytospin (Laborclin; cat. 620529).

### Airway lavage

This procedure was performed as previously described (Veras et al., 2020). Briefly, the airway fluid from 11 patients with COVID-19 patients was obtained by aspiration of the orotracheal tube. This fluid was mixed 1:1 with 0.1 M dithiothreitol (Thermo Fisher Scientific; cat. R0862), incubated for 15 min stirring every 5 min at 37°C. In the control group (n=8), the airway lavage was obtained by injecting sterile isotonic saline solution through a nasal fossa. The injected solution was mixed with nasal and nasopharyngeal secretions before being evacuated from the other nostril when it was collected directly in a sterile tube. The samples were centrifuged 750 g at 4°C for 10 min. The supernatants were used for measurement of NETs and the cells were fixed for immunostaining in coverslips with Poly-L-lysine solution 0.1% (Sigma-Aldrich; cat P8920).

### NETs quantification by MPO/DNA conjugates in picogreen assay

This procedure was performed as previously described (Czaikoski et al., 2016). Briefly, an antibody bound to a 96-well clear-bottom black plate captured the enzyme MPO (5 μg/ml;Abcam). According to the manufacturer’s instructions, the amount of DNA bound to the enzyme was quantified using the Quant-iT™ PicoGreen® kit (Invitrogen). The fluorescence intensity (excitation at 488 nm and emission at 525 nm) was quantified by a FlexStation 3 Microplate Reader (Molecular Devices, CA, USA). Neutrophils (10^6^ cells) obtained from COVID-19 severe patients or healthy controls in RPMI 1640 supplemented with 0.5% BSA were treated or not. The concentration of NETs in supernatants was determined and 4 h after infection NETs were determined in supernatants. In another context, neutrophils from healthy controls were treated or not 1 h before infection with SARS-CoV-2 (MOI = 1.0) and incubated for 4 h at 37°C. Next, NETs amounts were analyzed by picogreen assay. Furthermore, plasma or supernatants of airway fluid were incubated for 4 h at 37°C for determination of NETs.

### Human GSDMD ELISA Kit

GSDMD *in vitro* SimpleStep ELISA® (Enzyme-Linked Immunosorbent Assay) kit (ab272463) was used for the quantitative measurement of GSDMD in human plasma from COVID-19 moderate or severe and healthy control samples according to the manufacturer’s instructions. To perform the assay, samples or standards were added to the wells, followed by the antibody mix. TMB Development Solution was added and this reaction was then stopped by the addition of Stop Solution. The signal was measured at 450 nm.

### Immunofluorescence assay and confocal microscopy

Human and mice lung sections or isolated neutrophils were fixed with paraformaldehyde (4%). The samples were washed with PBS and blocked with 1% BSA, 22.5 mg/mL glycine in PBST (PBS + 0.1% Tween 20). The slides were overnight stained with the following antibodies: a) Mouse anti-myeloperoxidase (MPO, 2C7, Abcam, cat. ab25989, 1:500); B) Rabbit anti-GSDMD-NT (Abcam, cat. ab215203). Next, the samples were incubated with alpaca anti-mouse IgG Alexa Fluor 488 (Jackson ImmunoResearch, cat. 615-545-214, 1:1000) or alpaca anti-rabbit IgG Alexa Fluor 594 (Jackson ImmunoResearch, cat. 611-585-215, 1:1000) secondary antibodies for 30 minutes. The nuclei were stained with 4’,6-diamidino-2-phenylindole, dihydrochloride (DAPI, Life Technologies, cat. D1306, 1:1.000). The Axio Observer acquired images combined with the LSM 780 confocal microscope system (Carl Zeiss) at a 630x magnification. All images acquired were analyzed using Fiji by ImageJ. Finally, 10 fields per sample were randomically analyzed at the x and y focal plane.

### Cells, virus, and mock

To obtain the SARS-CoV-2 virus or the control (Mock) the SARS-CoV-2 Brazil/SPBR-02/2020 strain was isolated from a nasopharyngeal swab of the first confirmed Brazilian cases of COVID-19 and expanded on Vero-E6 (African green monkey kidney) cells. Vero-E6 cells were cultured in DMEM high glucose supplemented with 10% fetal bovine serum (FBS; HyClone, Logan,Utah) and 100 U/mL penicillin, and 1% μg/mL streptomycin (P/S Corning; cat. 30-002-CI), 1% glutamine (Corning; cat. 15718008), 1% hepes (Corning; cat.25-060-CI), and 1% fungizone (Gibco; cat. 15290-018) at 37°C in the 5% CO2 atmosphere. Experiments were performed after one passage in cell culture when Vero-E6 cells with DMEM plus 2% FBS in 150 cm^2^ surface area flasks were incubated at 37°C in 5% CO2 atmosphere. All procedures related to virus culture were handled at a biosafety level 3 (BSL3) multi-user facility, according to WHO guidelines. Virus titers were determined as the tissue culture infectious dose at 50% (TCID 50/mL). Virus stocks were kept in −80 °C ultra-freezers. The virus strain was sequenced to confirm the virus identity and its complete genome is publicly deposited (GenBank accession No. MT710714). Non-infected control cultures (mock) were prepared using pure non-supplemented DMEM as inoculum.

### Mouse infection and treatment

To evaluate the effects of Disulfiram *in vivo*, we infected the K18-hACE2 humanized mice (B6.Cg-Tg(K18-ACE2)2Prlmn/J) (McCray et al., 2007; Oladunni et al., 2020; Bao et al., 2020). K18-hACE2 mice were obtained from The Jackson Laboratory and were bred in the Centro de Criação de Animais Especiais (Ribeirão Preto Medical School/University of São Paulo). For the experimental infection, animals were transferred to the BSL3 facility. Eight-week-old male K18-hACE2 mice were infected with 2×10^4^ PFU of SARS-CoV-2 (in 40 μL) by the intranasal route as previously described (Oladunni et al, 2020). Twenty-four hours after the virus inoculation and once daily until day 5 post-infection (dpi), animals were treated with Disulfiram (50 mg/kg, i.p.) (n = 7) or vehicle (n = 7). Naive mice remained uninfected and untreated. On the 5 dpi, 6 h after the injection of Disulfiram or vehicle, animals were humanely euthanized for samples collection. All the experimental procedures were performed in accordance with the guide for the use of laboratory animals of the University of Sao Paulo and approved by the institutional ethics committee (066/2020).

### Drugs

To assess the pathways involved in the release of NETs, neutrophils were treated with GSDMD inhibitor - Disulfiram - (Tetraethylthiuram disulfide) - T24201 - (3, 10, or 30 μM); RNA polymerase inhibitor - TDF - (tenofovir disoproxil fumarate) – CAS 202138-50-9 - (10 μM); neutralizing anti-hACE2 antibody - αACE2 - RheaBiotech; Cat. IM- 0060 - (0.5 μg/ml); serine protease TMPRSS2 - camostat mesylate - Cat. SML0057 - (10 μM); pan-caspase inhibitor - Z-VAD-FMK - CAS187389-52-2 - (50 μM); caspase-1 Inhibitor I - CAS 143313-51-3 - (25 μM); or vehicle (DMSO, <1% v/v) 1 h before stimulation.

### Plaque Reduction Neutralization Test using disulfiram against SARS-CoV-2

A PRNT (Plaque Reduction Neutralization Test) was performed in VERO E6 cells on a cell-monolayer in 24-well plates to evaluate the effect of disulfiram on SARS-CoV-2 infection. A serial dilution of disulfiram was made in MEM medium, no FBS, using a 4-fold dilution factor, from 1mM to 0,0156 mM and then incubated 1 hour at 37ºC with approximately 90 PFU (Plaque Forming Units) of SARS-CoV-2. Cells were washed twice with a saline buffer (PBS), and the complex virus+disulfiram was added to the confluent monolayer. Plates were incubated for one hour at 37ºC to allow virus adsorption. Cells were washed with PBS twice, and a semi-solid carboxymethyl cellulose (CMC) medium (1.5% in DMEM) was added to the cells and incubated four days at 37ºC, 5% CO2. The overlay was removed, and cells fixed with formalin 10% buffer and then stained for 15 minutes in 1% crystal violet. PRNT50 value was calculated using a curve fitting method (Nonlinear dose-response regression) to a more precise result. The data are expressed as relative PFU (%) to that for the untreated virus-infected control cells, which was defined as 100%.

### Cytopathic effect of SARS-CoV-2 infected neutrophils on A549 and HUVEC cell lines by flow cytometry

Neutrophils were isolated from healthy controls (1×10^6^) incubated or not with disulfiram (10 μM) for 1 h. Next, were incubated or not with SARS-CoV-2 (MOI = 1.0) for 1 h. Then, the infected neutrophils were washed twice and co-cultured with Human alveolar basal epithelial (A549) or Human Umbilical Vein Endothelial Cells (Huvec) cell lines (2× 10^5^) previously stained with CellTrace™ Violet Cell Proliferation Kit (Thermo Fisher #C34557) following manufacturer protocols, for 24 h at 37°C. Subsequently, the cells were harvested and suspended in buffer containing 2% FBS in PBS for further evaluation of cell viability by flow cytometry through staining with Fixable and viability dye eFluor™ 780 (eBiosciente™) following manufacturer protocol (Veras et al., 2020). The acquisition of the cells was performed in a flow cytometer (FACS Verse) and the analyses were made using the FlowJo software (FlowCytometry Analysis Software v10).

### Cytokine assays (ELISA)

The quantification of cytokines in mouse lung was conducted by a commercial ELISA kit (R&D Systems) according to the manufacturer’s instructions. The individual sample’s optical density was measured at 450 nm using a spectrophotometer (Spectra Max-250, Molecular Devices, Sunnyvale, CA, USA).

### Western Blot

Neutrophils (3 × 10^6^) from blood were isolated by percoll density gradients as previously described (3) and were lysed with a boiling Laemmli buffer. Samples were loaded onto a 15% SDS-PAGE gel. Proteins were then transferred onto nitrocellulose membranes using Bio-Rad’s Trans-Blot Turbo, which were blocked using 5% nonfat dry milk in 1X TBS-T buffer for 1 h at room temperature. The membranes were incubated overnight at 4°C under mild agitation in 5% nonfat dry milk in 1X TBS-T buffer containing the following primary antibodies: mouse anti-Caspase-1 (p20) (mAb (Bally-1); Adipogen; AG-20B-0048-C100; 1:1000), mouse anti-Caspase-4 (MBL cat. M029-3; 1:1000), rabbit anti-GSDMD (Abcam, cat. ab215203; 1:1000), rabbit anti-α-actin (Sigma-Aldrich, cat. A2066; 1:5000), mouse anti-α-actin (Cell Signaling, cat. 3700; 1:1000), rabbit anti-RIG-I(D14G6) (Cell Signaling, cat. 3743S; 1:1000). Membranes were washed in 1x TBS-T and incubated with appropriate secondary HRP-conjugated antibodies diluted in 5% nonfat dry milk in 1X TBS-T buffer. Protein detection was done using an ECL™ Prime Western Blotting System (GE Healthcare) and an Amersham Imager 600 (GE Healthcare).

### Histological examination

Mice were euthanized 5 days post-SARS-CoV-2 infection. The lung tissue was harvested and fixed in 4% buffered formalin and posteriorly embedded in paraffin blocks. Sections (3 μm) were then stained with hematoxylin and eosin for histological examination. Images were acquired by DMI 6000B microscope (LeicaMicrosystems) at a 400x magnification. Histological evaluation was performed by a pathologist.

### Statistics

Statistical significance was determined by either two-tailed paired or unpaired Student t-test or one-way ANOVA followed by Tukey’s post hoc test. Absolute numbers and percentages were compared with Fisher’s exact test. Spearman’s rank-order correlation (r) was calculated to describe correlations. *P*<0.05 was considered statistically significant. Statistical analyses and graph plots were performed with GraphPad Prism 8.4.2 software.

## Data Availability

All data produced in the present study are available upon reasonable request to the authors

## Supplementary material

In supplementary figure 1, we showed that GSDMD expression is associated with NETs in airway fluid from COVID-19 severe patients. In supplementary figure 2, we showed the evaluation of the antiviral effect of disulfiram against SARS-CoV-2. In supplementary figure 3, we demonstrated that RIG-I is highly expressed in neutrophils from COVID-19 patients. In supplementary figure 4, are showing the gating strategy for flow cytometry analysis. In supplementary figure 5, we demonstrated that GSDMD inhibition reduces organ damage in a COVID-19 mouse model.

## Author contributions

Conception and design of the study: Silva CM, Wanderley CW, and Cunha FQ. Acquisition of data (provided animals, acquired and managed patients, provided facilities, etc.): Silva CM; Wanderley CW; Veras FP; Gonçalves AV; Lima MHF; Toller-Kawahisa JE; Gomes GF; Nascimento DC; Monteiro VV; Paiva IM; Almeida CJLR; Caetité DB; Silva JC; Rodrigues TS; Lopes MI; Bonjorno LP; Giannini MC; Amaral NB; Benatti MN; Damasceno LEA; Silva BM; Schneider AH; Castro IMS; Silva JCS; Vasconcelos AP; Gonçalves TT; Batah SS; Costa VF; Pontelli MC; Martins RB; Martins TV; Cebinelli GCM. Analysis and interpretation of data (e.g., statistical analysis, biostatistics, computational analysis): Silva CM; Wanderley CW; Nakaya HI. Writing, review, and/or revision of the manuscript: Silva CM; Wanderley CW; Leira LOS; Cunha LD; Arruda E; Fonseca BAL; Nakaya HI; Fabro AT; Oliveira RD; Zamboni DS; Louzada-Junior P; Cunha TM; Alves-Filho JC; Cunha FQ.

## Acknowledgments

We are grateful to Marcella Daruge Grando, Livia Maria C.S. Ambrósio, Muriel C.R.O. Berti, Basílica Botelho Muniz, and Juliana Trench Abumansur for technical assistance. This research was supported by Fundação de Amparo à Pesquisa do Estado de São Paulo grants (2013/08216-2 and 2020/05601-6), Conselho Nacional de Desenvolvimento Científico e Tecnológico and Coordenação de Aperfeiçoamento de Pessoal de Nível Superior grants.

## Competing interests

The authors declare that they have no competing interests.

## Funding

This work was supported by grants from the São Paulo Research Foundation (FAPESP) under grant agreement no. 2013/08216-2 (Center for Research in Inflammatory Diseases) and the Coordenação de Aperfeiçoamento de Pessoal de Nível Superior (CAPES).

## Supplementary Figures

**Supplementary figure 1.**
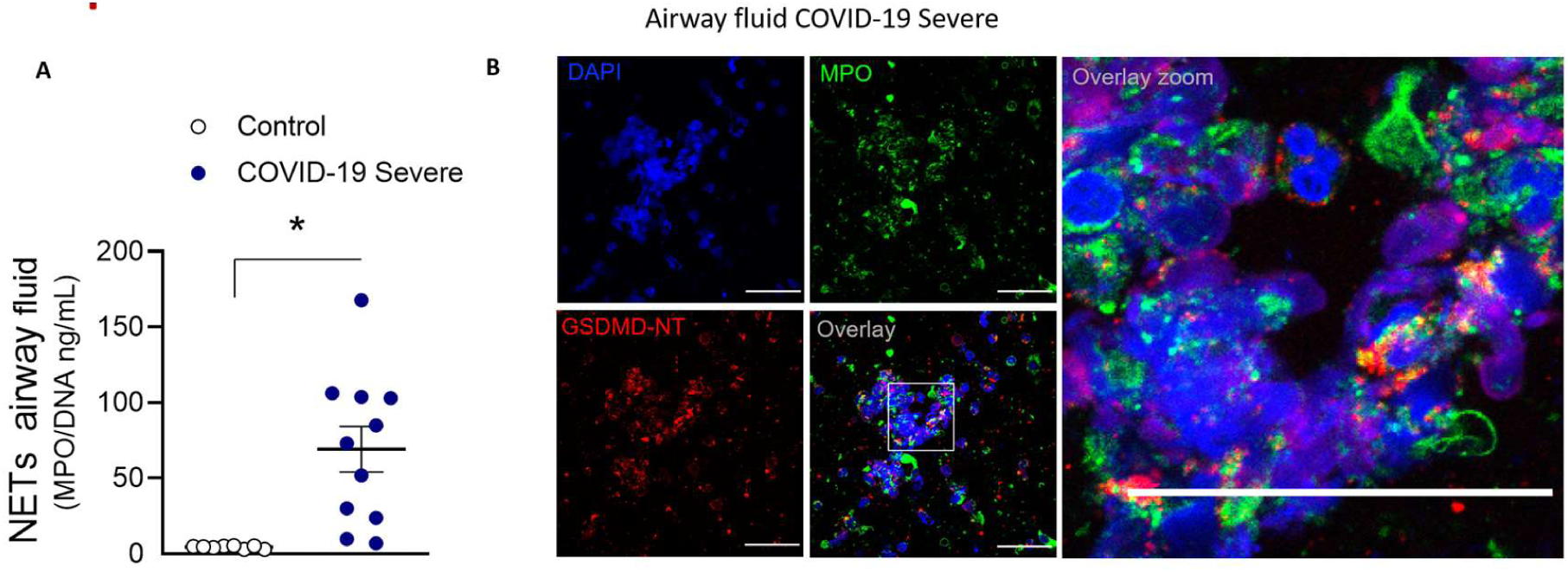
GSDMD expression is associated with NETs in airway fluid from COVID-19 severe patients. **(A)** NET quantification by picogreen assay with MPO-DNA in the airway fluid from COVID-19 patients (n=11) and in saline-induced airway fluids from healthy control (n=8). **(B)** Representative confocal analysis of GSDMD and NETs from the airway fluid of COVID-19 severe patients. Cells were stained for DNA (DAPI, blue), MPO (green), and GSDMD-NT (red). Scale bar indicates 50 μm. 4× digital zoom was performed in the inset white square. The data are expressed as mean ± SEM (*p<0.05; t-test in A).

**Supplementary figure 2.**
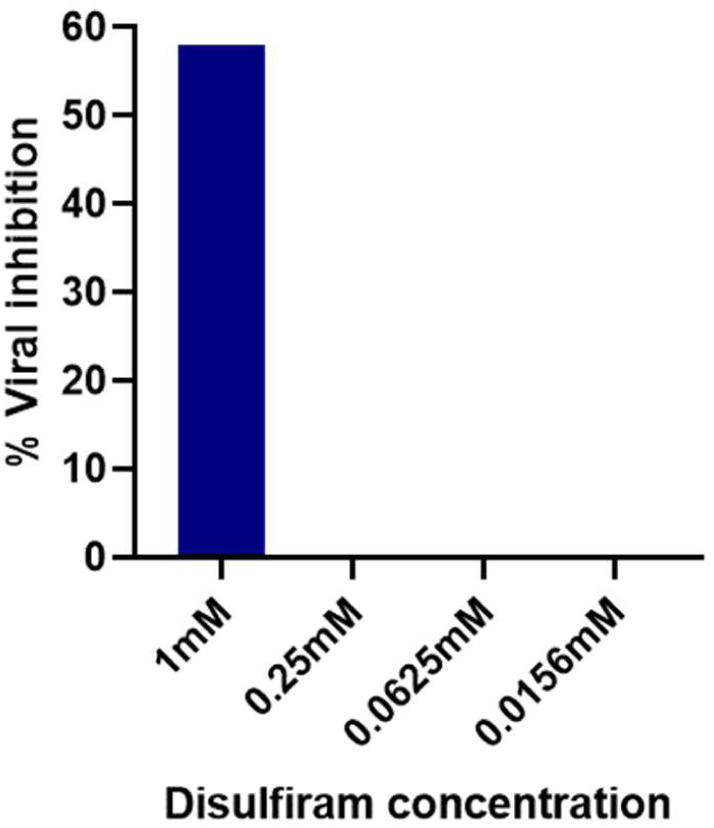
Evaluation of the antiviral effect of disulfiram against SARS-CoV-2. Vero E6 cells were treated with disulfiram (using a 4-fold serial dilution) and then incubated 1 hour at 37ºC with approximately 90 PFU (Plaque Forming Units) of SARS-CoV-2. Four days after infection, viral release to the media was measured by the plaque-forming unit (PFU) assay.

**Supplementary figure 3.**
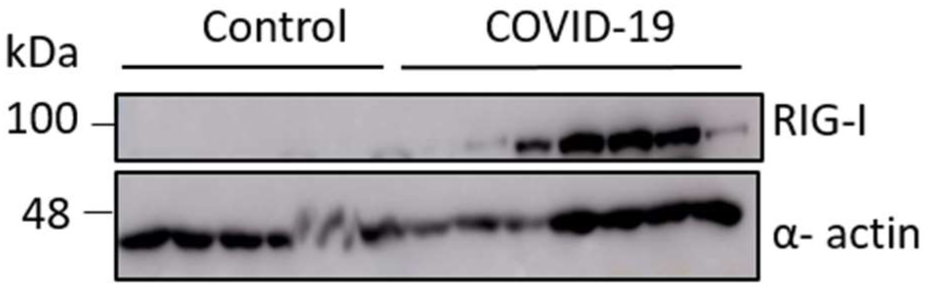
RIG-I is highly expressed in neutrophils from COVID-19 patients. Neutrophils were isolated from healthy controls (n=7) and COVID-19 patients (n=8). The neutrophil lysates were harvested for immunoblot analysis of RIG-1. The α-actin was used as a loading control.

**Supplementary figure 4.**
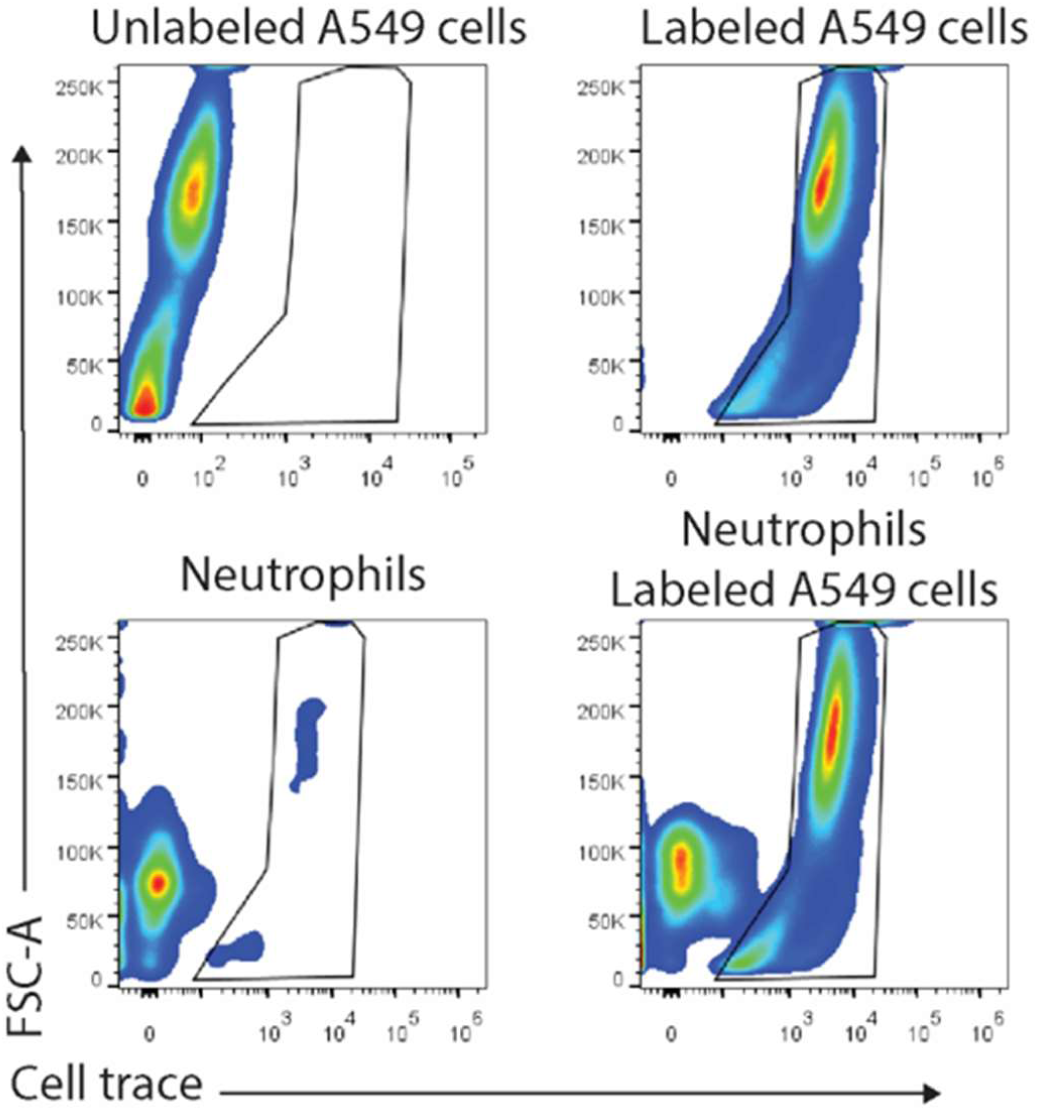
Gating strategy for flow cytometry analysis. Blood isolated neutrophils (10^6^ cells) from healthy donors, pretreated, or not, with disulfiram (30 μM) were incubated, or not, with SARS-CoV-2. After 1 h, these cells were washed twice and co-cultured with lung epithelial cells (A549, 2 × 10^5^ cells) or endothelial cells (HUVEC, 2 × 10^5^ cells) previously stained with viability dye for 24 h at 37°C. Gating strategy for flow cytometry analysis of A549 or HUVEC viability.

**Supplementary figure 5.**
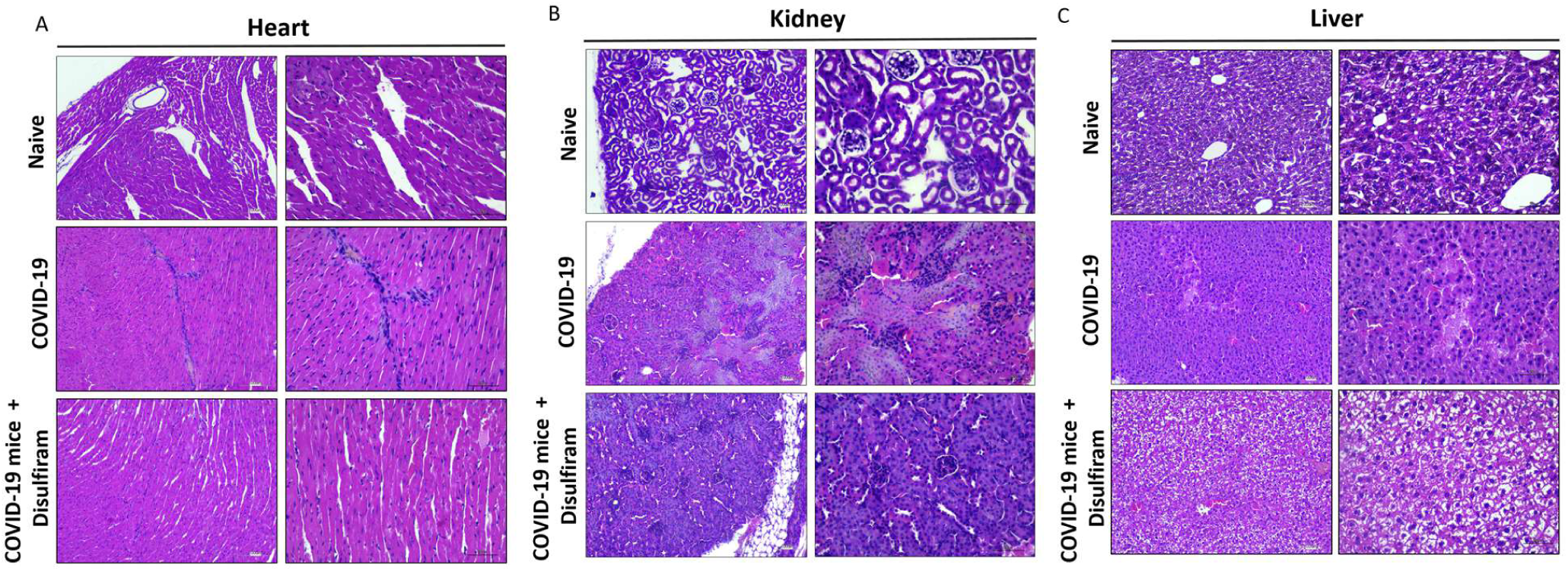
GSDMD inhibition reduces organ damage in a COVID-19 mouse model. ACE-2 humanized mice were infected with SARS-CoV-2 (2×10^4^) and after 24 h we treated mice with disulfiram (50 mg/kg, i.p. 1x per day, during 5 days) or vehicle. **(A)** Representative images of the histological staining of the **(A)** heart, **(B)** kidney, and **(C)** Liver sections performed 5 days post-SARS-CoV-2 infection are shown at 200× magnification and at 400× magnification. The data are representative of groups, each including 5-7 animals per group.

## Notes

### Competing Interest Statement

The authors have declared no competing interest.

### Funding Statement

This work was supported by grants from the Sao Paulo Research Foundation (FAPESP) under grant agreement no. 2013/08216-2 (Center for Research in Inflammatory Diseases) and the Coordenacao de Aperfeicoamento de Pessoal de Nivel Superior (CAPES).

### Author Declarations

Ethics committee/IRB of National Ethics Committee, Brazil (30248420.9.0000.5440) gave ethical approval for this work

## References

Ackermann, M., Anders, H. J., Bilyy, R., Bowlin, G. L., Daniel, C., De Lorenzo, R., Egeblad, M., Henneck, T., Hidalgo, A., Hoffmann, M., Hohberger, B., Kanthi, Y., Kaplan, M. J., Knight, J. S., Knopf, J., Kolaczkowska, E., Kubes, P., Leppkes, M., Mahajan, A., Manfredi, A. A., … Herrmann, M. 2021. Patients with COVID-19: in the dark-NETs of neutrophils. Cell death and differentiation, 28(11), 3125–3139. https://doi.org/10.1038/s41418-021-00805-z

Bao, L., Deng, W., Huang, B., Gao, H., Liu, J., Ren, L., Wei, Q., Yu, P., Xu, Y., Qi, F., Qu, Y., Li, F., Lv, Q., Wang, W., Xue, J., Gong, S., Liu, M., Wang, G., Wang, S., Song, Z., … Qin, C. 2020. The pathogenicity of SARS-CoV-2 in hACE2 transgenic mice. Nature, 583(7818), 830–833. https://doi.org/10.1038/s41586-020-2312-y

Brinkmann, V., Reichard, U., Goosmann, C., Fauler, B., Uhlemann, Y., Weiss, D. S., Weinrauch, Y., & Zychlinsky, A. 2004. Neutrophil extracellular traps kill bacteria. Science (New York, N.Y.), 303(5663), 1532–1535. https://doi.org/10.1126/science.1092385

Broz, P., Pelegrín, P., & Shao, F. 2020. The gasdermins, a protein family executing cell death and inflammation. Nature reviews. Immunology, 20(3), 143–157. https://doi.org/10.1038/s41577-019-0228-2

Carmona-Rivera, C., Carlucci, P. M., Goel, R. R., James, E., Brooks, S. R., Rims, C., Hoffmann, V., Fox, D. A., Buckner, J. H., & Kaplan, M. J. 2020. Neutrophil extracellular traps mediate articular cartilage damage and enhance cartilage component immunogenicity in rheumatoid arthritis. JCI insight, 5(13), e139388. https://doi.org/10.1172/jci.insight.139388

Chen, K. W., Monteleone, M., Boucher, D., Sollberger, G., Ramnath, D., Condon, N. D., von Pein, J. B., Broz, P., Sweet, M. J., & Schroder, K. 2018. Noncanonical inflammasome signaling elicits gasdermin D-dependent neutrophil extracellular traps. Science immunology, 3(26), eaar6676. https://doi.org/10.1126/sciimmunol.aar6676

Clososki, G.C., Soldi, R.A., Guaratini, T., Martins, R.B., Costa, C.S., Arruda, E., Lopes, N.P. 2020. Tenofovir Disoproxil Fumarate: New Chemical Developments and Encouraging, 31:1552–1556. https://doi.org/10.21577/0103-5053.20200106

Czaikoski, P. G., Mota, J. M., Nascimento, D. C., Sônego, F., Castanheira, F. V., Melo, P. H., Scortegagna, G. T., Silva, R. L., Barroso-Sousa, R., Souto, F. O., Pazin-Filho, A., Figueiredo, F., Alves-Filho, J. C., & Cunha, F. Q. 2016. Neutrophil Extracellular Traps Induce Organ Damage during Experimental and Clinical Sepsis. PloS one, 11(2), e0148142. https://doi.org/10.1371/journal.pone.0148142

Duarte-Neto, A. N., Monteiro, R., Johnsson, J., Cunha, M., Pour, S. Z., Saraiva, A. C., Ho, Y. L., da Silva, L., Mauad, T., Zanotto, P., Saldiva, P., de Oliveira, I., & Dolhnikoff, M. 2019. Ultrasound-guided minimally invasive autopsy as a tool for rapid post-mortem diagnosis in the 2018 Sao Paulo yellow fever epidemic: Correlation with conventional autopsy. PLoS neglected tropical diseases, 13(7), e0007625. https://doi.org/10.1371/journal.pntd.0007625

Elion, D. L., & Cook, R. S. 2018. Harnessing RIG-I and intrinsic immunity in the tumor microenvironment for therapeutic cancer treatment. Oncotarget, 9(48), 29007–29017. https://doi.org/10.18632/oncotarget.25626

Fillmore, N., Bell, S., Shen, C., Nguyen, V., La, J., Dubreuil, M., Strymish, J., Brophy, M., Mehta, G., Wu, H., Lieberman, J., Do, N., & Sander, C. 2021. Disulfiram use is associated with lower risk of COVID-19: A retrospective cohort study. PloS one, 16(10), e0259061. https://doi.org/10.1371/journal.pone.0259061

Funchal, G. A., Jaeger, N., Czepielewski, R. S., Machado, M. S., Muraro, S. P., Stein, R. T., Bonorino, C. B., & Porto, B. N. 2015. Respiratory syncytial virus fusion protein promotes TLR-4-dependent neutrophil extracellular trap formation by human neutrophils. PloS one, 10(4), e0124082. https://doi.org/10.1371/journal.pone.0124082

Hao, Y., Hao, S., Andersen-Nissen, E., Mauck, W. M., 3rd, Zheng, S., Butler, A., Lee, M. J., Wilk, A. J., Darby, C., Zager, M., Hoffman, P., Stoeckius, M., Papalexi, E., Mimitou, E. P., Jain, J., Srivastava, A., Stuart, T., Fleming, L. M., Yeung, B., Rogers, A. J., … Satija, R. 2021. Integrated analysis of multimodal single-cell data. Cell, 184(13), 3573–3587.e29. https://doi.org/10.1016/j.cell.2021.04.048

Hiroki, C. H., Toller-Kawahisa, J. E., Fumagalli, M. J., Colon, D. F., Figueiredo, L., Fonseca, B., Franca, R., & Cunha, F. Q. 2020. Neutrophil Extracellular Traps Effectively Control Acute Chikungunya Virus Infection. Frontiers in immunology, 10, 3108. https://doi.org/10.3389/fimmu.2019.03108

Hoffmann, M., Kleine-Weber, H., Schroeder, S., Krüger, N., Herrler, T., Erichsen, S., Schiergens, T. S., Herrler, G., Wu, N. H., Nitsche, A., Müller, M. A., Drosten, C., & Pöhlmann, S. 2020. SARS-CoV-2 Cell Entry Depends on ACE2 and TMPRSS2 and Is Blocked by a Clinically Proven Protease Inhibitor. Cell, 181(2), 271–280.e8. https://doi.org/10.1016/j.cell.2020.02.052

Hu, J. J., Liu, X., Xia, S., Zhang, Z., Zhang, Y., Zhao, J., Ruan, J., Luo, X., Lou, X., Bai, Y., Wang, J., Hollingsworth, L. R., Magupalli, V. G., Zhao, L., Luo, H. R., Kim, J., Lieberman, J., & Wu, H. 2020. FDA-approved disulfiram inhibits pyroptosis by blocking gasdermin D pore formation. Nature immunology, 21(7), 736–745. https://doi.org/10.1038/s41590-020-0669-6

Jin, Y. H., Cai, L., Cheng, Z. S., Cheng, H., Deng, T., Fan, Y. P., Fang, C., Huang, D., Huang, L. Q., Huang, Q., Han, Y., Hu, B., Hu, F., Li, B. H., Li, Y. R., Liang, K., Lin, L. K., Luo, L. S., Ma, J., Ma, L. L., et al. 2020. A rapid advice guideline for the diagnosis and treatment of 2019 novel coronavirus (2019-nCoV) infected pneumonia (standard version). Military Medical Research, 7(1), 4. https://doi.org/10.1186/s40779-020-0233-6

Kambara, H., Liu, F., Zhang, X., Liu, P., Bajrami, B., Teng, Y., Zhao, L., Zhou, S., Yu, H., Zhou, W., Silberstein, L. E., Cheng, T., Han, M., Xu, Y., & Luo, H. R. 2018. Gasdermin D Exerts Anti-inflammatory Effects by Promoting Neutrophil Death. Cell reports, 22(11), 2924–2936. https://doi.org/10.1016/j.celrep.2018.02.067

Knight, J. S., Luo, W., O’Dell, A. A., Yalavarthi, S., Zhao, W., Subramanian, V., Guo, C., Grenn, R. C., Thompson, P. R., Eitzman, D. T., & Kaplan, M. J. 2014. Peptidylarginine deiminase inhibition reduces vascular damage and modulates innate immune responses in murine models of atherosclerosis. Circulation research, 114(6), 947–956. https://doi.org/10.1161/CIRCRESAHA.114.303312

Koppaka, V., Thompson, D. C., Chen, Y., Ellermann, M., Nicolaou, K. C., Juvonen, R. O., Petersen, D., Deitrich, R. A., Hurley, T. D., & Vasiliou, V. 2012. Aldehyde dehydrogenase inhibitors: a comprehensive review of the pharmacology, mechanism of action, substrate specificity, and clinical application. Pharmacological reviews, 64(3), 520–539. https://doi.org/10.1124/pr.111.005538

Leppkes, M., Knopf, J., Naschberger, E., Lindemann, A., Singh, J., Herrmann, I., Stürzl, M., Staats, L., Mahajan, A., Schauer, C., Kremer, A. N., Völkl, S., Amann, K., Evert, K., Falkeis, C., Wehrfritz, A., Rieker, R. J., Hartmann, A., Kremer, A. E., Neurath, M. F., … Herrmann, M. 2020. Vascular occlusion by neutrophil extracellular traps in COVID-19. EBioMedicine, 58, 102925. https://doi.org/10.1016/j.ebiom.2020.10292

Lex, A., Gehlenborg, N., Strobelt, H., Vuillemot, R., & Pfister, H. 2014. UpSet: Visualization of Intersecting Sets. IEEE transactions on visualization and computer graphics, 20(12), 1983–1992. https://doi.org/10.1109/TVCG.2014.2346248

Liao, M., Liu, Y., Yuan, J., Wen, Y., Xu, G., Zhao, J., Cheng, L., Li, J., Wang, X., Wang, F., Liu, L., Amit, I., Zhang, S., & Zhang, Z. 2020. Single-cell landscape of bronchoalveolar immune cells in patients with COVID-19. Nature medicine, 26(6), 842–844. https://doi.org/10.1038/s41591-020-0901-9

Lopes, M. I., Bonjorno, L. P., Giannini, M. C., Amaral, N. B., Menezes, P. I., Dib, S. M., Gigante, S. L., Benatti, M. N., Rezek, U. C., Emrich-Filho, L. L., Sousa, B., Almeida, S., Luppino Assad, R., Veras, F. P., Schneider, A., Rodrigues, T. S., Leiria, L., Cunha, L. D., Alves-Filho, J. C., Cunha, T. M., … Oliveira, R. 2021. Beneficial effects of colchicine for moderate to severe COVID-19: a randomised, double-blinded, placebo-controlled clinical trial. RMD open, 7(1), e001455. https://doi.org/10.1136/rmdopen-2020-001455

McCray, P. B., Jr, Pewe, L., Wohlford-Lenane, C., Hickey, M., Manzel, L., Shi, L., Netland, J., Jia, H. P., Halabi, C., Sigmund, C. D., Meyerholz, D. K., Kirby, P., Look, D. C., & Perlman, S. 2007. Lethal infection of K18-hACE2 mice infected with severe acute respiratory syndrome coronavirus. Journal of virology, 81(2), 813–821. https://doi.org/10.1128/JVI.02012-06

Oladunni, F. S., Park, J. G., Pino, P. A., Gonzalez, O., Akhter, A., Allué-Guardia, A., Olmo-Fontánez, A., Gautam, S., Garcia-Vilanova, A., Ye, C., Chiem, K., Headley, C., Dwivedi, V., Parodi, L. M., Alfson, K. J., Staples, H. M., Schami, A., Garcia, J. I., Whigham, A., Platt, R. N., 2nd, … Torrelles, J. B. 2020. Lethality of SARS-CoV-2 infection in K18 human angiotensin-converting enzyme 2 transgenic mice. Nature communications, 11(1), 6122. https://doi.org/10.1038/s41467-020-19891-7

Papayannopoulos V. 2018. Neutrophil extracellular traps in immunity and disease. Nature reviews. Immunology, 18(2), 134–147. https://doi.org/10.1038/nri.2017.105

Radermecker, C., Detrembleur, N., Guiot, J., Cavalier, E., Henket, M., d’Emal, C., Vanwinge, C., Cataldo, D., Oury, C., Delvenne, P., & Marichal, T. 2020. Neutrophil extracellular traps infiltrate the lung airway, interstitial, and vascular compartments in severe COVID-19. The Journal of experimental medicine, 217(12), e20201012. https://doi.org/10.1084/jem.20201012

Rintahaka, J., Wiik, D., Kovanen, P. E., Alenius, H., & Matikainen, S. 2008. Cytosolic antiviral RNA recognition pathway activates caspases 1 and 3. Journal of immunology (Baltimore, Md. : 1950), 180(3), 1749–1757. https://doi.org/10.4049/jimmunol.180.3.1749

Rodrigues, T. S., de Sá, K., Ishimoto, A. Y., Becerra, A., Oliveira, S., Almeida, L., Gonçalves, A. V., Perucello, D. B., Andrade, W. A., Castro, R., Veras, F. P., Toller-Kawahisa, J. E., Nascimento, D. C., de Lima, M., Silva, C., Caetite, D. B., Martins, R. B., Castro, I. A., Pontelli, M. C., de Barros, F. C., … Zamboni, D. S. 2021. Inflammasomes are activated in response to SARS-CoV-2 infection and are associated with COVID-19 severity in patients. The Journal of experimental medicine, 218(3), e20201707. https://doi.org/10.1084/jem.20201707

Saitoh, T., Komano, J., Saitoh, Y., Misawa, T., Takahama, M., Kozaki, T., Uehata, T., Iwasaki, H., Omori, H., Yamaoka, S., Yamamoto, N., & Akira, S. 2012. Neutrophil extracellular traps mediate a host defense response to human immunodeficiency virus-1. Cell host & microbe, 12(1), 109–116. https://doi.org/10.1016/j.chom.2012.05.015

Shi, J., Zhao, Y., Wang, K., Shi, X., Wang, Y., Huang, H., Zhuang, Y., Cai, T., Wang, F., & Shao, F. 2015. Cleavage of GSDMD by inflammatory caspases determines pyroptotic cell death. Nature, 526(7575), 660–665. https://doi.org/10.1038/nature15514

Shi, S., Qin, M., Shen, B., Cai, Y., Liu, T., Yang, F., Gong, W., Liu, X., Liang, J., Zhao, Q., Huang, H., Yang, B., & Huang, C. 2020. Association of Cardiac Injury With Mortality in Hospitalized Patients With COVID-19 in Wuhan, China. JAMA cardiology, 5(7), 802–810. https://doi.org/10.1001/jamacardio.2020.0950

Silva, C., Wanderley, C., Veras, F. P., Sonego, F., Nascimento, D. C., Gonçalves, A. V., Martins, T. V., Cólon, D. F., Borges, V. F., Brauer, V. S., Damasceno, L., Silva, K. P., Toller-Kawahisa, J. E., Batah, S. S., Souza, A., Monteiro, V. S., Oliveira, A., Donate, P. B., Zoppi, D., Borges, M. C., … Cunha, F. Q. 2021. Gasdermin D inhibition prevents multiple organ dysfunction during sepsis by blocking NET formation. Blood, 138(25), 2702–2713. https://doi.org/10.1182/blood.2021011525

Sollberger, G., Choidas, A., Burn, G. L., Habenberger, P., Di Lucrezia, R., Kordes, S., Menninger, S., Eickhoff, J., Nussbaumer, P., Klebl, B., Krüger, R., Herzig, A., & Zychlinsky, A. 2018. Gasdermin D plays a vital role in the generation of neutrophil extracellular traps. Science immunology, 3(26), eaar6689. https://doi.org/10.1126/sciimmunol.aar6689

Veras, F. P., Pontelli, M. C., Silva, C. M., Toller-Kawahisa, J. E., de Lima, M., Nascimento, D. C., Schneider, A. H., Caetité, D., Tavares, L. A., Paiva, I. M., Rosales, R., Colón, D., Martins, R., Castro, I. A., Almeida, G. M., Lopes, M., Benatti, M. N., Bonjorno, L. P., Giannini, M. C., Luppino-Assad, R., … Cunha, F. Q. 2020. SARS-CoV-2-triggered neutrophil extracellular traps mediate COVID-19 pathology. The Journal of experimental medicine, 217(12), e20201129. https://doi.org/10.1084/jem.20201129

Wright, C., & Moore, R. D. 1990. Disulfiram treatment of alcoholism. The American journal of medicine, 88(6), 647–655. https://doi.org/10.1016/0002-9343(90)90534-k

Wu, Z., & McGoogan, J. M. 2020. Characteristics of and Important Lessons From the Coronavirus Disease 2019 (COVID-19) Outbreak in China: Summary of a Report of 72 314 Cases From the Chinese Center for Disease Control and Prevention. JAMA, 323(13), 1239–1242. https://doi.org/10.1001/jama.2020.2648

Yamada, T., Sato, S., Sotoyama, Y., Orba, Y., Sawa, H., Yamauchi, H., Sasaki, M., & Takaoka, A. 2021. RIG-I triggers a signaling-abortive anti-SARS-CoV-2 defense in human lung cells. Nature immunology, 22(7), 820–828. https://doi.org/10.1038/s41590-021-00942-0

Zeng, H., Ma, Y., Zhou, Z., Liu, W., Huang, P., Jiang, M., Liu, Q., Chen, P., Luo, H., & Chen, Y. 2021. Spectrum and Clinical Characteristics of Symptomatic and Asymptomatic Coronavirus Disease 2019 (COVID-19) With and WithoutPneumonia. Frontiers in medicine, 8, 645651. https://doi.org/10.3389/fmed.2021.645651

Zhang, C., Shi, L., & Wang, F. S. 2020. Liver injury in COVID-19: management and challenges. The lancet. Gastroenterology & hepatology, 5(5), 428–430. https://doi.org/10.1016/S2468-1253(20)30057-1

Zhou, F., Yu, T., Du, R., Fan, G., Liu, Y., Liu, Z., Xiang, J., Wang, Y., Song, B., Gu, X., Guan, L., Wei, Y., Li, H., Wu, X., Xu, J., Tu, S., Zhang, Y., Chen, H., & Cao, B. 2020. Clinical course and risk factors for mortality of adult inpatients with COVID-19 in Wuhan, China: a retrospective cohort study. Lancet (London, England), 395(10229), 1054–1062. https://doi.org/10.1016/S0140-6736(20)30566-3

